# Pathogen exposure misclassification can bias association signals in GWAS of infectious diseases when using population-based common controls

**DOI:** 10.1101/2022.07.14.22276656

**Authors:** Dylan Duchen, Candelaria Vergara, Chloe L. Thio, Prosenjit Kundu, Nilanjan Chatterjee, David L. Thomas, Genevieve L. Wojcik, Priya Duggal

## Abstract

Genome-wide association studies (GWAS) have been performed to identify host genetic factors for a range of phenotypes, including for infectious diseases. The use of population-based common controls from biobanks and extensive consortiums is a valuable resource to increase sample sizes in the identification of associated loci with minimal additional expense. Non-differential misclassification of the outcome has been reported when the controls are not well-characterized, which often attenuates the true effect size. However, for infectious diseases the comparison of cases to population-based common controls regardless of pathogen exposure can also result in selection bias. Through simulated comparisons of pathogen exposed cases and population-based common controls, we demonstrate that not accounting for pathogen exposure can result in biased effect estimates and spurious genome-wide significant signals. Further, the observed association can be distorted depending upon strength of the association between a locus and pathogen exposure and the prevalence of pathogen exposure. We also used a real data example from the hepatitis C virus (HCV) genetic consortium comparing HCV spontaneous clearance to persistent infection with both well characterized controls, and population-based common controls from the UK Biobank. We find biased effect estimates for known HCV clearance-associated loci and potentially spurious HCV clearance-associations. These findings suggest that the choice of controls is especially important for infectious diseases or outcomes that are conditional upon environmental exposures.

## INTRODUCTION

Genome-wide association studies (GWAS) are focused on identifying genetic associations with health outcomes. This has been successfully accomplished using well-characterized cases and controls often from epidemiologic cohort or case-control studies.^1^ More recently, GWAS have utilized biobanks and consortiums to expand sample populations. This can include comparing well-characterized cases to phenotypically uncharacterized or population-based common controls which has led to the identification of novel associations with low to modest effect size s.^2–9^ Concerns related to the use of common controls include not adequately accounting for population substructure and the likelihood of non-differential misclassification across the outcome (i.e. some proportion of the controls have developed the outcome of interest) which can often attenuate true associations.^2,6,8–10^

When studying infectious disease, it is key that an individual is exposed to a pathogen (i.e. virus, bacteria, protozoa) before they can develop disease. Host genetics and immunity along with pathogen genetics, co-infections, co-morbidities, age, and sex are known to explain some of the heterogeneity in disease outcomes. Thus, it is critical to account for pathogen exposure because unexposed individuals are never at risk of developing the outcome. This can be achieved through antibody or antigen testing or obtaining documented history of exposure (i.e. vaccine, contact tracing). While it is assumed that the internal validity of case-control studies, including GWAS, is maintained by the characterization of both the genetic exposure and the phenotypic outcome for every study participant, it is also assumed that cases and controls are at risk of developing the outcome. For infectious disease studies involving population-based common controls, this presumption is not always true and can result in differential misclassification of pathogen exposure between cases and controls.^11,12^ Whether epidemiological factors that influence pathogen exposure (i.e. increased transmission in certain populations or occupations, routes of transmission, or comorbidities) can subsequently result in spurious genetic association signals due to this misclassification is not fully explored. In this study, we performed simulations comparing cases to well-characterized controls with known exposures and to population-based common controls with unknown exposures to identify if external variables associated with pathogen exposure can induce spurious associations. We also used empirical data to compare the genetic association results from a GWAS performed to identify host loci associated with recovery from hepatitis C virus (HCV) infection using either known HCV-exposed and persistently infected controls or population-based common controls from the UK Biobank (UKB).

## SUBJECTS AND METHODS

### Simulations to characterize pathogen exposure-associated selection bias

The directed acyclic graph (DAG) depicted in **Figure 1** provides a graphical representation of how comparing exposed cases to population-based common controls differs from comparisons made to well-characterized (pathogen-exposed) controls. For loci associated with pathogen exposure (e.g., a causal locus for a risk factor of pathogen exposure) unrelated to the outcome of interest, if unequivocally pathogen exposed cases are compared to population-based common controls regardless of exposure status, differential misclassification of pathogen exposure can induce selection bias, resulting in spurious associations between the outcome and the loci associated with pathogen exposure.

**Figure 1:**
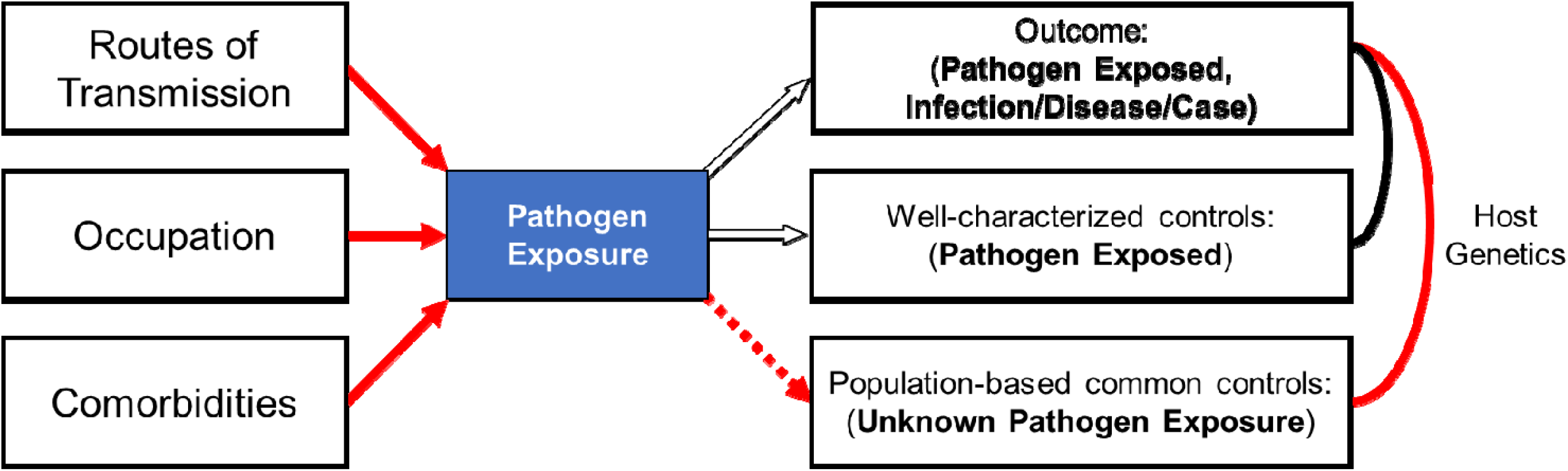
Conceptual framework of pathogen exposure-linked selection bias due to the differential misclassification of pathogen exposure and the use of population-based common controls. GWAS comparing unequivocally exposed cases to controls regardless of pathogen exposure can result in spurious (false positive) associations between loci not associated with the outcome if the loci are associated with pathogen exposure. The comparison of universally exposed cases to semi-exposed controls (host genetics, red line comparison) results in differential misclassification of pathogen exposure, introducing associations between pathogen exposure (and risk factors for pathogen exposure) and the outcome. No spurious association is expected when pathogen exposed cases are compared to pathogen exposed well-characterized controls (host genetics, black line comparison). Arrows dictate the direction of hypothesized causal effects. Hollow arrows reflect conditional relationships between exposure and outcome/case selection. The dashed arrow reflects an association induced via the differential misclassification of pathogen exposure. Red arrows highlight paths involved with pathogen exposure-linked selection bias, which results in observed associations between risk factors of pathogen exposure (and their linked loci) and an outcome of interest when cases are compared to population-based common controls of unknown pathogen exposure status.

How strongly pathogen exposure is associated with case status determines the strength of this bias (i.e., the dashed red line reflecting the degree of differential misclassification in pathogen exposure in **Figure 1**). Simulations were performed to assess whether the association between a non-outcome associated SNP can become spuriously associated with the outcome due to a relationship with pathogen exposure. Whether the prevalence of a pathogen contributes to this bias was also determined by assessing the degree to which inflated effect estimates for the outcome~SNP relationship were observed across the various simulation scenarios. Simulations were performed using the lavaan package (v0.6-8) in R as it allows for the control of statistical associations between simulated variables.^13,14^ The phenotypic data and SNP genotypes were simulated for a large cohort (N=1,000,000).

#### Case and control definitions

**Cases** were defined as individuals with a known pathogen exposure who developed the clinical outcome. **‘Well-characterized controls’** were defined as individuals with a known pathogen exposure event but did not develop the clinical outcome. To investigate the effects of differential misclassification of pathogen exposure in the absence of non-differential misclassification of the outcome, ‘**population-based common controls’** were defined as any individual who did not develop the clinical outcome, regardless of pathogen exposure. In each simulated cohort, a fixed number of cases, well-characterized controls, and population-based common controls were randomly sampled from individuals meeting the inclusion criteria. In each simulated cohort, unless otherwise stated, the prevalence of pathogen exposure was set at 25%. Cases were selected from the exposed population, of which 50% had the observed clinical outcome and the remainder defined as well-characterized controls. As population-based common controls were selected from the entire population, excluding cases, a proportion of the population-based controls would be expected to have a simulated pathogen exposure.

#### Simulated variables

Additional simulated variables included a single nucleotide polymorphism (SNP, coded as 0,1, 2 minor alleles) and a variable associated with pathogen exposure (U1 which could represent routes of transmission, occupation, comorbidities, etc.). To explore the effect of confounding within a locus completely unrelated the outcome of interest, neither the SNP nor the U1 variable were simulated to be associated with the outcome. For all simulated cohorts, the SNP had a fixed minor allele frequency (MAF) of 15% and a fixed SNP~U1 association (β) of 0.1, whereby each additional SNP allele was associated with an increase of 10% of U1. Similar effect sizes have been observed for markers associated with other complex diseases.^15–19^ The SNP was simulated with a MAF of 15% and in Hardy Weinberg Equilibrium (HWE). The effect estimates for associations between simulated variables were confirmed via logistic and linear model-based regressions using the speedglm package in R.^13,14^

#### Simulation scenario 1 – Effects of a pathogen exposure-specific variable

We simulated 500,000 replicates of N=1,000,000 individuals for each of the following relationships between U1 and pathogen exposure: no association (β=0, OR=1), a moderate association (β=log(1.2), OR=1.2), or a strong association (β=log(2), OR=2). Simulations were performed assuming the prevalence of pathogen exposure was 25% and 50% of exposed individuals had the outcome (**Table 1**).

**Table 1:** Simulation scenario parameters. For each scenario, the parameter of interest, the fixed values, and the sample sizes of each simulation is listed.

| Simulation |  | Scenario-specific parameter | Fixed Values |  |  |  |
| --- | --- | --- | --- | --- | --- | --- |
| | | | Outcome: Post-exposure | Pathogen Exposure Prevalence | $\beta$: U1~Pathogen Exposure | Sample size: Cases (N):Controls (N) |
| Scenario 1a: Cases vs. Population-Based Controls | | $\beta$: U1~Pathogen Exposure | 50% | 25% | $\log(1)$, $\log(1.2)$, $\log(2)$ | 20,000 : 20,000<br>20,000 : 200,000 |
| Scenario 1b: Cases vs. Well-Characterized Controls |  |  |  |  |  | 20:000 : 20:000 |
| Scenario 2a: Cases vs. Population-Based Controls | | Pathogen Exposure Prevalence | 50% | 5%, 25%, 50%, 75%, 100% | $\log(1.2)$<br>$\log(2.0)$ | 20,000 : 20,000<br>20,000 : 200,000 |
| Scenario 2b: Cases vs. Well-Characterized Controls |  |  |  |  |  | 20:000 : 20:000 |

#### Simulation scenario 2 – Effects of the prevalence of pathogen exposure

We simulated 500,000 replicates of N=1,000,000 individuals for each of the following pathogen exposure prevalences: 5%, 25%, 50%, 75%, or 100%. Simulations were performed assuming either a moderate (β=log(1.2), OR=1.2) or a strong association (β=log(2), OR=2) between U1 and pathogen exposure and 50% of exposed individuals had the outcome (**Table 1**).

#### Simulated genetic associations

In each simulated replicate, 20,000 cases, 20,000 well-characterized controls, and 20,000 population-based controls were randomly selected and included in logistic regressions to quantify the ‘outcome of interest’~SNP association. Regressions compared cases to population-based controls (scenarios 1a and 2a) or well-characterized controls (scenarios 1b and 2b). To simulate a realistic use-case of GWAS involving many common controls, we re-performed the above simulations and regressions for both scenarios comparing 20,000 cases to 200,000 population-based controls. Regressions were performed in R using the Rfast package.^20^

To test whether the effect sizes of the SNP~outcome associations (β_SNP_) were significantly different when cases were compared to population-based or well-characterized controls, we derived a modified Z statistic, appropriate for the comparison of regression coefficients,^21^ reflecting the change in average scenario-specific beta estimates for each parameter between comparisons involving population-based controls and well-characterized controls. The standard error term used to estimate each Z score was defined as the mean standard error across each scenario’s parameter-specific set of simulations. This conservative test was chosen due to the extreme number of comparisons (e.g., with two logistic regressions for each of the parameter-specific 500,000 replicates, 13 million regressions were performed when comparing 20,000 cases to 20,000 population-based controls).

As a null association for the ‘outcome of interest’~SNP relationship was simulated for each cohort, any non-null observed ‘outcome of interest’~SNP association reflect spurious signals induced via the scenario-specific selection of case and controls. We calculated the proportion of cohorts with ‘outcome of interest’~SNP associations that reached the conservative but widely accepted genome-wide significance threshold of P≤5×10^−8^.^22^ To determine whether the prevalence of pathogen exposure was associated with the magnitude of these spurious signals, we performed linear regressions to obtain the magnitude and direction of the average beta estimates obtained from each set of pathogen exposure prevalence parameter-specific simulations. Measures of heterogeneity were estimated using the meta R package for each set of parameter-specific simulations to confirm independent yet equivalent cohorts were simulated (**Table S1**).^23^ Additional simulations were considered and are included in the supplementary methods.

### HCV GWAS using cases and well characterized controls or population-based controls

#### Well characterized cases and controls from the HCV Extended Genetics Consortium

We leveraged data from The HCV Extended Genetic Consortium (HCV Consortium) which includes 1,869 individuals of African ancestry, 1,739 individuals of European ancestry, and 486 individuals of Hispanic ancestry who passed GWAS-related quality control metrics,^24^ as previously described.^25–27^ We only used individuals of European ancestry in this study (tightly clustered with the 1000 Genomes Project (1000G) Northern European population from Utah (CEU) and had an admixture estimate of at least 90% European ancestry, determined via fastSTRUCTURE).^28^ Of the 1,739 European ancestry HCV consortium individuals, 702 were individuals with serological evidence of spontaneous clearance from HCV infection (cases) and 1,037 were defined as individuals with persistent HCV infection (“well-characterized controls”).^25^ Of these individuals, 16% were living with HIV and 31% were female.^25–27^

The UK Biobank (UKB) is a large population-level cohort including 500,000 volunteers recruited from across the UK.^29^ ‘Population-based controls’ were unrelated individuals from the UKB who were identified to be of similar genetic ancestry to HCV Consortium individuals of European ancestry. For these individuals, we had no information on HCV infection status or previous exposure to HCV.

#### Selection of samples for analysis

##### Ancestry analysis of the combined HCV Consortium-UKB cohort

Linkage disequilibrium (LD)-based pruning was used to iteratively remove variants in LD (r^2^ >0.2) via a sliding 500k base-pair-wide window prior to performing principal components analysis (PCA) in the combined HCV Consortium-UKB cohort. This LD-based pruning was performed on all genetic markers with missingness <1.5% and MAF >2.5%, after excluding regions of long-range linkage disequilibrium.^30^ Given the extreme sample size of the combined HCV-UKB cohorts, FastPCA was used.^31,32^ LD-based pruning and PCA was carried out using the R package SNPRelate.^33^

##### Selection of ancestry-matched UKB population-based controls

Out of 487,409 UKB individuals with genetic data available, 407,192 UKB participants who passed previously described QC metrics were included within our analyses.^29,34,35^ We limited UKB population-based controls to 370,702 ancestry-matched individuals genetically similar to the European individuals from the HCV Consortium (**Figures S1-S2**).^36^ Briefly, genetic ancestry-based matching of cases and controls was accomplished through multiple iterations of PCA. Initially, the UKB participants whose top three PCs were within six standard deviations of the mean PC values estimated from HCV Consortium individuals were retained for analysis (N=383,724). A second PCA filtering step, using the top twenty PCs, excluded any UKB individual with estimated eigenvalues 5% higher or lower than the range of PC-specific values from HCV Consortium individuals. A total of 370,702 ancestry-matched UKB controls were identified.

#### Selection of markers for analysis

##### HCV Consortium

Out of an initial 661,397 directly genotyped markers mapped to the GRCh37/hg19 reference sequence, a total of 656,340 markers had MAF >1%, variant-level call rates >97%, were in Hardy-Weinberg Equilibrium (HWE) based on HWE exact tests (P>1×10^−5^), and passed the ‘McCarthy Group Tools’ Haplotype Reference Consortium (HRC) imputation preparation pipeline (https://www.well.ox.ac.uk/~wrayner/tools/).^37^ No allele frequency difference threshold between this database and the HCV Consortium cohort individuals was used to remove genotyped markers. Imputation was performed using the HRC (version r1.1, 2016) European reference panel via the Michigan Imputation Server and phased using Eagle (v2.4).^38,39^ Markers with imputation quality R^2^ values > 0.3 were retained and markers which were not bi-allelic or had a genotyping rate < 97% or MAF < 1% were removed. Additionally, imputed markers with variant-level INFO scores <90% or HWE P<5×10^−7^ were removed. A total of 6,468,618 imputed markers passed these QC metrics.

##### UK Biobank

A total of 670,739 directly genotyped autosomal QC-passed markers were used for imputation, as previously described.^29^ Phasing was performed with SHAPEIT3 and imputation carried out using IMPUTE4.^29,40,41^ UKB imputation involved a combination of the Haplotype Reference Consortium (HRC), the UK10K haplotype reference, and the 1000 Genomes phase 3 reference panels.^29^ To be considered a potential match for any QC-passed HCV Consortium-derived marker, the imputed UKB variant was required to have INFO score >90%, MAF>1%, and HWE P>1×10^−7^. A total of to 93,095,623 imputed UKB variants met this quality control threshold and were considered for analysis.

##### Combination of the HCV Consortium and UKB datasets

Imputed markers in the HCV consortium were matched to UKB QC-passed imputed markers using position and allele information. A total of 6,025,969 markers were shared across the ancestry-matched UKB and HCV Consortium databases. A total of 6,009,835 markers were shared across the combined HCV Consortium and a set of 364,308 UKB controls who passed a more stringent set of sample-level genetic heterozygosity-focused QC metrics than those performed by the UKB consortium, as described below and in the **Supplementary Methods**.

#### Selection of different subsets of UKB population-based controls for exploratory GWAS

Additional exploratory GWAS using different subsets of UKB controls were performed to determine whether the observed association between certain genetic loci and HCV clearance was driven by mismatched fine-scale ancestry, excess sample-level genetic heterozygosity, or unaccounted for population structure due to the extreme number of population-based UKB controls (**Figures S3-S8**). Other exploratory GWAS assessed differential enrichment for certain epidemiological factors relative to the population based common controls. This included a GWAS comparing the set of HCV clearance cases from non-hemophilia-focused cohorts (N=513) to a more precisely matched case-control cohort of UKB controls (N=4,908) (**Figure S9**). A GWAS comparing all the individuals from the HCV Consortium (N=1,739) to the European ancestry-matched population-based controls from the UKB (N=370,702) was also performed (**Figure S10**). Each exploratory GWAS occurred after performing all sample-level and imputed marker-level QC described above.

### Statistical analysis

#### HCV GWAS using cases and well characterized controls or population-based controls

##### Association of cases and well characterized controls

Genetic associations between the 702 cases and 1,037 well-characterized controls of the HCV Consortium were performed via logistic regression under an additive genetic model using PLINK,^32,42^ as the lack of any significant sample size imbalance between cases and controls or intensive computational resources needed to perform these genetic associations did not warrant a more complex association approach. Covariates for this GWAS included sex and the first twenty principal components, estimated from the HCV Consortium dataset after linkage disequilibrium-based pruning.

##### Association of cases and population-based controls

To account for the severe sample size imbalance between cases and controls, and to optimize computational performance at extreme differences in sample sizes, associations between the 702 cases and 370,702 ancestry-matched population-based controls were performed under an additive genetic model using REGENIE with the saddle point approximation (SPA) test ^3,43,44^ While REGENIE partially accounts for population stratification, the top twenty PCs estimated from the combined HCV-UKB cohort were included as fixed effect covariates to ensure differences due to genetic ancestry or population structure among UKB individuals of European ancestry were accounted for (**Figure S2**).^45,46^ Sex was included as a covariate.

We utilized a genome-wide significance threshold of P≤5×10^−8^.^22^ After performing the association testing, to minimize the risk of bias due to differences across genotyping arrays or imputation panels between our cases and controls among putative HCV clearance-associated loci, a MAF-based filter was used to exclude markers suggestively associated with HCV clearance (P≤5×10^−5^) using genome-wide allele frequency data for the gnomAD non-Finnish European population downloaded in September 2021, using the gnomAD v2.1.1 release.^10^ Any suggestively associated marker with a MAF>10% among gnomAD non-Finnish Europeans was excluded if the MAF among UKB controls and gnomAD non-Finnish Europeans differed in absolute terms by at least 5%. Similarly, any suggestively associated marker with a MAF<10% in gnomAD non-Finnish Europeans was excluded if the relative difference between the gnomAD-derived MAF and UKB MAF was greater than 25%.

Manhattan and quantile-quantile plots for each GWAS were generated using the ggfastman or qqman packages in R (**Figure S14**).^47^ A table of all suggestively associated markers (P<5×10^−5^) for the ancestry-matched UKB controls GWAS can be found within the supplemental material (**Supplementary Table 2**).

We performed Cochran’s *Q* test of heterogeneity using METAL to determine whether the results obtained from the GWAS of cases vs. population-based controls from the UKB differed significantly from the GWAS of cases vs. well-characterized controls from the HCV consortium (**Figure S11**).^48,49^

## RESULTS

### Simulations to characterize pathogen exposure-associated selection bias

Simulations were performed using a simplified version of the framework presented in **Figure 1**. Briefly, the association between a SNP not associated with outcome but associated with pathogen exposure via the variable ‘U1’, was compared between pathogen exposed cases and pathogen exposed (well-characterized) or population-based common controls.

First, we simulated whether the presence and magnitude of the U1~’pathogen exposure’ relationship affects the association between a SNP linked to U1 and the outcome of interest (**Table 1**). To provide a baseline for further explorations, the first model was simulated to have no association between U1 and pathogen exposure (OR=1). As expected, we found no association between the SNP and outcome, regardless of the choice of well-characterized or population-based controls or whether cases were compared to 20,000 population-based common controls or 200,000 population-based common controls (P=1) (**Figure 2**). For all simulation scenarios, no replicates comparing cases and well-characterized pathogen exposed controls resulted in spurious ‘outcome of interest’~SNP associations (**Table 2**).

**Table 2:** Proportion of spurious associations and odds ratios across scenario-specific simulated cohorts. For each scenario, the parameter of interest, the fixed values, and the sample sizes of each simulation is listed. OR: odds ratio.

|  |  |  |  | 20,000 Cases vs. 20,000 Common Controls |  | 20,000 Cases vs. 200,000 Common Controls |  |
| --- | --- | --- | --- | --- | --- | --- | --- |
| Scenario | Controls | Parameter Value | U1~Pathogen Exposure | OR (Mean) | Spurious Associations N (%) | OR (Mean) | Spurious Associations N (%) |
| 1 | Pathogen exposed | U1~Pathogen Exposure | OR=1 | 1 | 0 | 1 | 0 |
|  |  |  | OR=1.2 | 1 | 0 | 1 | 0 |
|  |  |  | OR=2 | 1 | 0 | 1 | 0 |
|  | Population-based | U1~Pathogen Exposure | OR=1 | 1 | 0 | 1 | 0 |
|  |  |  | OR=1.2 | 1.04 | 159 (0.03) | 1.04 | 1,792 (0.36) |
|  |  |  | OR=2 | 1.13 | 410,503 (82.1) | 1.13 | 499,666 (99.93) |
| 2 | Pathogen exposed | Pathogen Exposure Prevalence: 5% | OR=1.2 | 1 | 0 (0) | 1 | 0 (0) |
|  | Population-based |  |  | 1.06 | 2,830 (0.57) | 1.06 | 36,815 (7.36) |
|  | Pathogen exposed |  | OR=2 | 1 | 0 (0) | 1 | 0 (0) |
|  | Population-based |  |  | 1.2 | 499,962 (99.99) | 1.197 | 500,000 (100) |
|  | Pathogen exposed | Pathogen Exposure Prevalence: 25% | OR=1.2 | 1 | 0 (0) | 1 | 0 (0) |
|  | Population-based |  |  | 1.04 | 153 (0.03) | 1.04 | 1,749 (0.35) |
|  | Pathogen exposed |  | OR=2 | 1 | 0 (0) | 1 | 0 (0) |
|  | Population-based |  |  | 1.13 | 407,864 (81.57) | 1.132 | 499,647 (99.93) |
|  | Pathogen exposed | Pathogen Exposure Prevalence: 50% | OR=1.2 | 1 | 0 (0) | 1 | 0 (0) |
|  | Population-based |  |  | 1.03 | 13 (<0.01) | 1.03 | 136 (0.03) |
|  | Pathogen exposed |  | OR=2 | 1 | 0 (0) | 1 | 0 (0) |
|  | Population-based |  |  | 1.095 | 101,203 (20.24) | 1.095 | 399,122 (79.82) |
|  | Pathogen exposed | Pathogen Exposure Prevalence: 75% | OR=1.2 | 1 | 0 (0) | 1 | 0 (0) |
|  | Population-based |  |  | 1.019 | 3 (<0.01) | 1.019 | 10 (<0.01) |
|  | Pathogen exposed |  | OR=2 | 1 | 0 (0) | 1 | 0 (0) |
|  | Population-based |  |  | 1.06 | 2,725 (0.55) | 1.059 | 35,680 (7.14) |
|  | Pathogen exposed | Pathogen Exposure Prevalence: 100% | OR=1.2 | 1 | 0 (0) | 1 | 0 (0) |
|  | Population-based |  |  | 1 | 1 (<0.01) | 1 | 0 (0) |
|  | Pathogen exposed |  | OR=2 | 1 | 0 (0) | 1 | 0 (0) |
|  | Population-based |  |  | 1 | 1 (<0.01) | 1 | 0 (0) |

**Figure 2:**
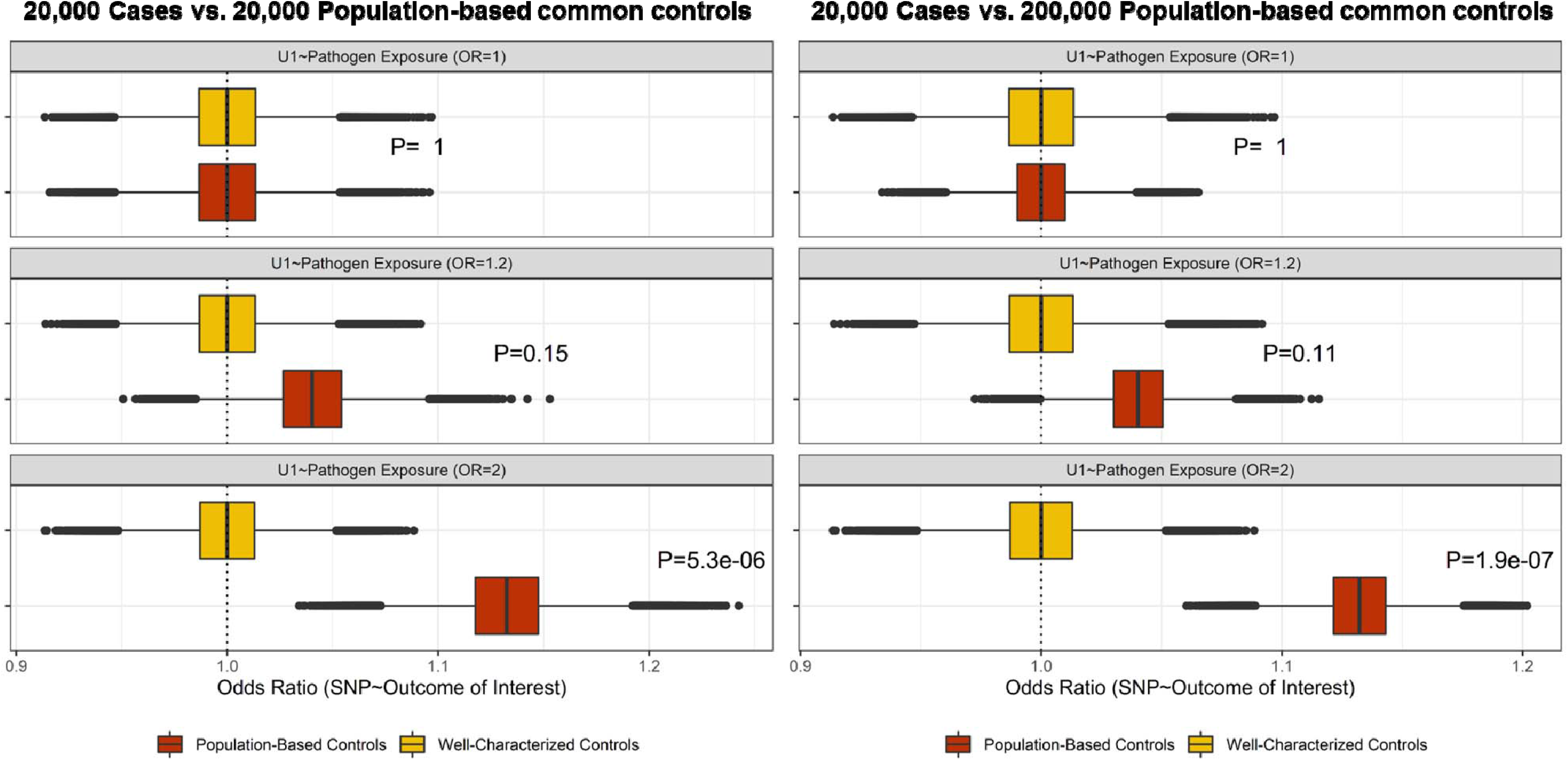
Distribution of observed odds ratios for a non-outcome associated locus across varying U1~Pathogen exposure associations. Comparisons involved 20,000 cases and equal numbers of well-characterized or population-based controls (left) or 20,000 cases and equal numbers of well-characterized and 200,000 population-based controls (right). Boxplots reflect distribution of odds ratios obtained comparing cases to population-based (red) or well-characterized controls (yellow). Reported P-values derived from a Z score based on the difference between averaged beta estimates when using population-based controls vs. well-characterized controls.

For cohorts with a moderate association between U1~’pathogen exposure’ (OR=1.2) there was an observed but non-significant inflation for comparisons involving 20,000 population based common controls compared to 20,000 well-characterized controls (P=0.15), based on the Z score estimate derived using the average effect estimates obtained when cases were compared to population-based or well-characterized controls. Additionally, a small number of replicates (N=159/500,000) had spurious genome-wide significant associations when cases were compared to population-based common controls. When the association between U1~’population exposure’ was simulated to be stronger (OR=2), significantly inflated odds ratios were observed for comparisons involving 20,000 population-based common controls (P=5.3×10^−6^) (**Figure 2**). For these simulations, 82.1% of replicates (N=410,503/500,000) had spurious associations (P<5×10^−8^) when cases were compared to 20,000 population-based common controls (**Table 2)**.

To investigate a scenario closer to current practice, we simulated unbalanced datasets with 20,000 cases and 200,000 population-based controls. In these simulations the reduced variance due to increased statistical power across SNP ~’outcome of interest’ estimates resulted in similarly inflated but higher proportions of spurious genome-wide significant replicates compared to replicates involving 20,000 population-based common controls for both moderate (N=1,792/500,000, P=0.11) and strong (N=499,666/500,000, P= 1.9 × 10^−7^) ‘U1~pathogen exposure’ association scenarios (**Figure 2, Table 2)**.

In the second set of simulations we varied the pathogen exposure prevalence from 5% to 100% to consider different infectious disease prevalence scenarios. All prevalence values except for universal exposure (100%) resulted in inflated effect estimates for comparisons involving population-based common controls when assuming either moderate (OR=1.2) or strong (OR=2) U1~pathogen exposure relationships, although only the strong exposure was significant (**Figure 3, Figure S12**). Using 20,000 cases vs. 200,000 population-based controls we observed increased spurious associations for rare pathogen exposure prevalence (5%, N=36,815/500,000) which decreased to 0 replicates with spurious associations as the pathogen exposure approached 100% (or all exposed). This was present for both moderate and strong U1~pathogen exposure associations (**Table 2**), suggesting any observed bias was related to the differential misclassification of pathogen exposure when cases were compared to population-based controls. For comparisons involving 20,000 cases and 20,000 population-based controls, a single spurious replicate was observed for the scenario involving 100% pathogen exposure prevalence (**Table 2**).

**Figure 3:**
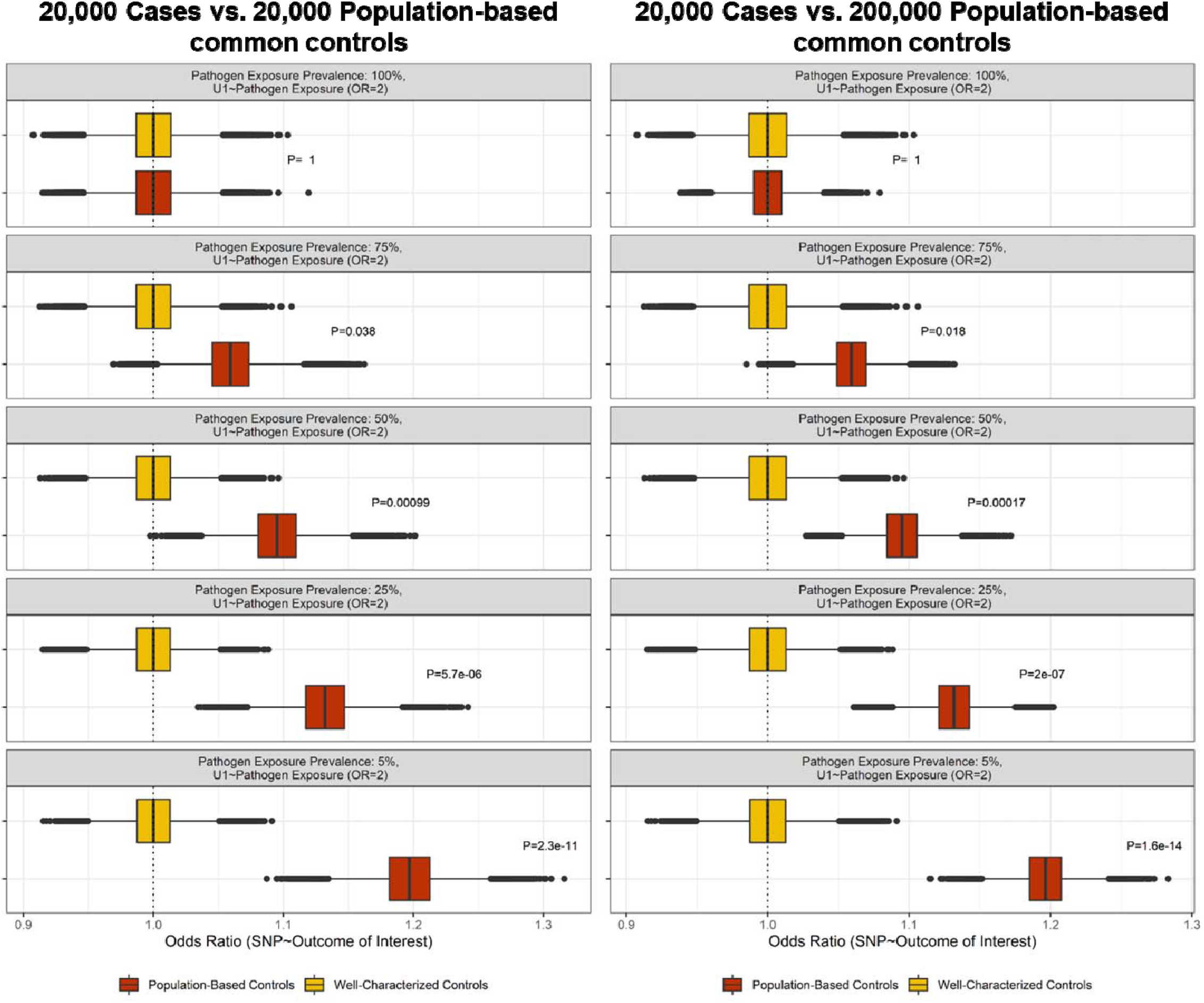
Distribution of observed odds ratios for a non-outcome strongly associated locus across varying pathogen exposure prevalence. Comparisons involved 20,000 cases and equal numbers of well-characterized or population-based controls (left) or 20,000 cases and equal numbers of well-characterized and 200,000 population-based controls (right) assuming a strong U1~’Pathogen exposure’ relationship. Boxplots reflect distribution of odds ratios comparing cases to population-based (red) or well-characterized controls (yellow). Reported P-values derived from a Z score based on the difference between averaged beta estimates when using population-based controls vs. well-characterized controls.

We also evaluated if the number of cases altered the magnitude of any inflated effect estimates since infectious disease studies are often much smaller than other complex disease studies due to availability and collection of samples. When assuming a strong U1~pathogen exposure relationship and fixing other parameters to those utilized in the first scenario simulations, no observable difference in the mean inflation of the outcome~SNP association (OR~1.13) was observed for comparisons between varying numbers of cases: 500, 1,000, 5,000, 10,000 and 20,000 and 200,000 population-based controls (**Figure S13**), suggesting that the bias exists regardless of case sample size.

### HCV GWAS using cases and well-characterized controls or population-based controls

To assess the real-world consequences of differential pathogen exposure misclassification in GWAS of infectious disease, we explored the effects of control definitions using a previously-published GWAS of HCV spontaneous clearance versus persistence.^25^ All cases and controls included within this previous GWAS were unequivocally exposed to HCV and the outcome of interest focused on a specific clinical sequela of infection, HCV clearance. We compared the results of this HCV clearance GWAS with the well-characterized (persistently HCV infected) controls to the same cases compared to population-based common controls from the UKB. The GWAS comparing HCV clearance cases to persistently infected individuals replicated previously published loci.^27,50–52^

#### Replication of known HCV clearance-associated loci

In the GWAS using population-based UKB controls (702 cases vs. 370,702 population-based controls), the two primary HCV clearance-associated loci within the human leukocyte antigen (HLA) region (*HLA-DQB1*) and *IFNL3* locus were replicated (P<5×10^−8^; **Figure 4**). However, the effect sizes of these markers were biased towards the null compared to the GWAS using well-characterized controls (**Table S3**). The genome-wide significant *IFNL3* locus markers had odds ratios in the range of 0.37-0.45 when cases were compared to well-characterized controls, while there were odds ratios of 0.56-0.63 for the population-based UKB controls comparison. The difference in effect size for each of these markers was significant (Cochran’s Q test, P<0.05, **Figure S11**), including the chromosome 19 marker in **Table 3**, and consistent with bias towards the null due to non-differential outcome misclassification from using population-based controls.

**Table 3:** Known HCV clearance associated loci. Results of the association for the top HCV clearance-associated markers within the *HLA-DQB1* and *IFNL3* loci from the GWAS comparing cases with well-characterized controls from the HCV Consortium or ancestry matched population-based common UKB controls. Measures of association and frequency of the effect allele for each locus are provided for each analysis. OR: Odds Ratio. 95% CI: 95% confidence interval of the OR. EAF: Effect allele frequency.

| Analyzed groups | Chromosome 6<br>*HLA-DQB1*, rs9275241 |  |  |  |  | Chromosome 19<br>*IFNL3*, rs11881222 |  |  |  |  |
| --- | --- | --- | --- | --- | --- | --- | --- | --- | --- | --- |
|  | OR | 95% CI | P value | Cases<br>(EAF, G) | Controls<br>(EAF, G) | OR | 95% CI | P value | Cases<br>(EAF, G) | Controls<br>(EAF, G) |
| Cases vs. well-characterized controls (HCV Persistence) | 0.614 | 0.532-0.708 | $1.90 \times 10^{-11}$ | 39.46% | 51.49% | 0.424 | 0.358-0.503 | $4.06 \times 10^{-23}$ | 20.09% | 36.31% |
| Cases vs. Ancestry-matched population-based controls (UKB) | 0.691 | 0.620-0.772 | $4.35 \times 10^{-11}$ | 39.46% | 52.12% | 0.609 | 0.541-0.686 | $4.04 \times 10^{-16}$ | 20.09% | 28.15% |

**Figure 4:**
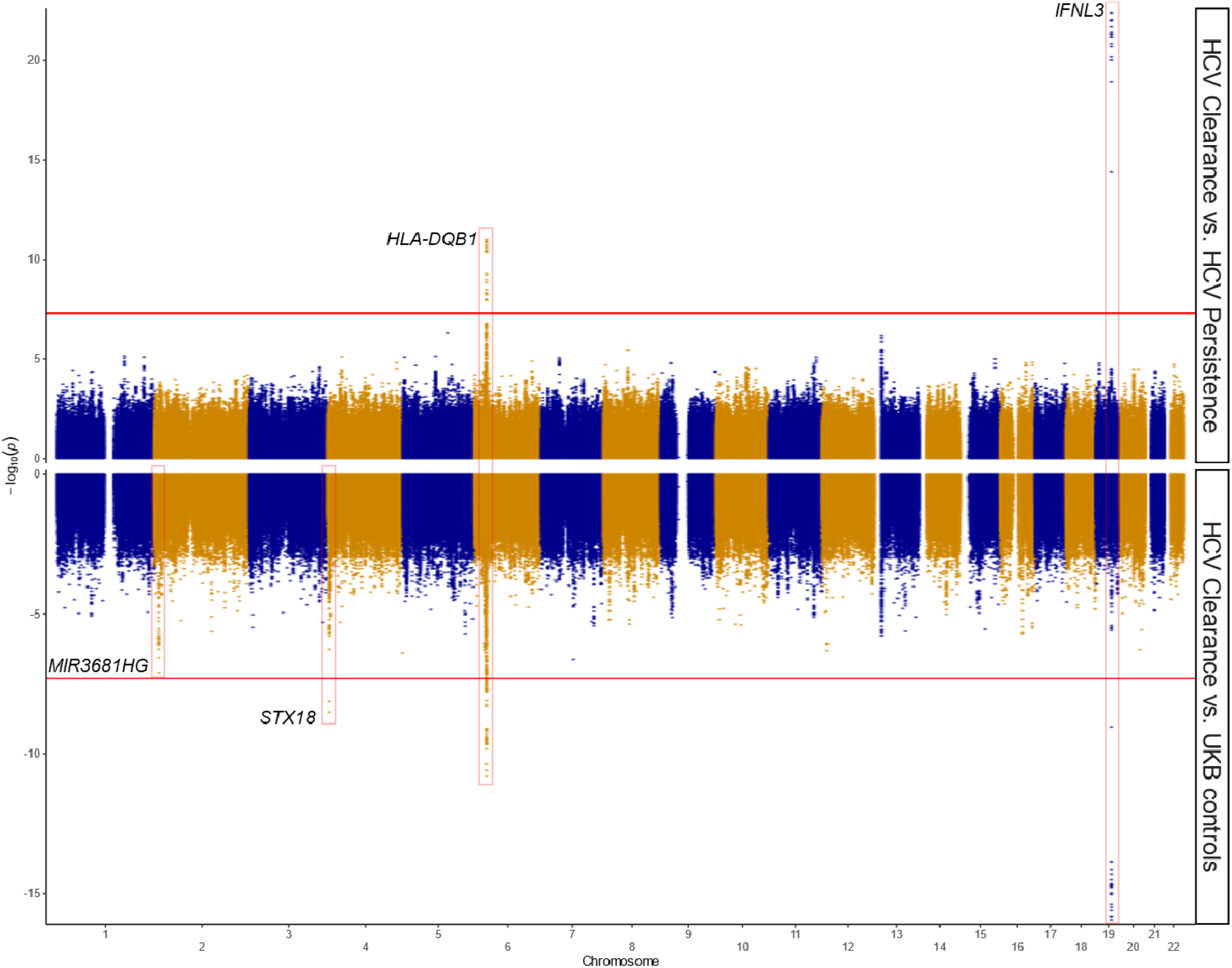
Manhattan plot of the GWAS comparing HCV clearance cases to persistently infected well-characterized controls (top) and ancestry-matched population-based UKB controls (bottom). The top panel reflects results from a GWAS performed comparing HCV clearance individuals (N=702) to individuals persistently infected with HCV from the HCV Consortium (N=1,037). The bottom panel reflects the results of a GWAS performed comparing the same HCV clearance cases to ancestry-matched population-based common controls from the UKB (N=370,702). The same set of genome-wide markers were used in each GWAS (6,025,969 markers). Points reflects the P values for each marker across the genome, ordered along the X axis by chromosomal position. The Y axis reflects the −log (P value), with the most significantly associated markers farthest away from 0 and outside the genome-wide significance threshold (P<5×10^−8^), indicated by the red lines. Loci of interest are highlighted and annotated with their nearest/overlapping gene, with novel HCV clearance-associated loci highlighted in the bottom panel.

#### Use of population-based controls identifies previously unknown HCV clearance-associated loci

In addition to the replication of known HCV clearance-associated loci, we identified two novel loci using population-based common controls. The first locus was on chromosome 4 within an intron of *syntaxin 18* (*STX18*) (rs58612183, MAF_UKB_=6.3%, OR=1.93, P=3.01×10^−9^, **Figure 4**) but had a reduced OR and was not significant in the GWAS involving well-characterized controls (MAF_Persistence_=9.5%, OR=1.22, P=0.084). Interestingly, the minor allele frequency in the control groups differed but was within the European range of 6-10% as reported in ensemble showing a north-south gradient.^53^ In the GWAS comparing all HCV European ancestry individuals from the HCV Consortium (persistence and clearance, N=1,739) to population-based UKB controls a genome-wide significant association was identified for this locus (P=3.9×10^−10^) suggesting that this locus is not HCV clearance-specific but reflects any HCV infection (or membership in the HCV Consortium) compared to population controls with unknown viral exposure. The signal in *STX18* remained associated with HCV clearance in additional exploratory GWAS except for the associations excluding cases known to have hemophilia and their ancestry-matched UKB controls (P=8.55×10^−4^, OR=1.47) (**Table 4**).

**Table 4:** Novel HCV clearance associated chromosome 4 locus. Results of the association for the top HCV clearance-associated marker within the *STX18* locus from the GWAS with cases vs. ancestry-matched population-based UKB controls. Measures of association and frequency of the effect allele (C) are also reported for the analysis of all cases vs all well-characterized controls GWAS and a matched case-control cohort GWAS excluding HCV clearance cases with hemophilia and their matched population-based controls. OR: Odds Ratio. 95% CI: 95% confidence interval of the OR. EAF: Effect allele frequency.

| Analyzed Groups | Chromosome 4:<br>*STX18*, rs58612183 |  |  |  |  |
| --- | --- | --- | --- | --- | --- |
|  | OR | 95% CI | P value | Clearance<br>(EAF, C) | Controls<br>(EAF, C) |
| Cases vs. well-characterized controls<br>(HCV Persistence) | 1.22 | 0.97-1.54 | $8.40 \times 10^{-2}$ | 10.97% | 9.50% |
| Cases vs. Ancestry-matched population-based<br>controls (UKB) | 1.93 | 1.56-2.41 | $3.01 \times 10^{-9}$ | 10.97% | 6.33% |
| No Hemophiliacs: Cases vs. Ancestry-matched<br>population-based controls (UKB, Matched 1:10) | 1.47 | 1.17-1.84 | $8.55 \times 10^{-4}$ | 9.55% | 6.65% |

The second locus was on chromosome 2 within the long noncoding RNA (lncRNA) *MIR3681HG* (top marker rs10803744, MAF_UKB_=12.5%, OR=1.57, P=7.99×10^−8^). Interestingly, the allele frequency differs between HCV clearance (MAF=17%) and HCV persistent (MAF=13%) individuals (OR=1.42, P=4.06×10^−4^) with European general populations matching the HCV persistent (ensemble MAF 11-15%).^53^ While some of the exploratory GWAS efforts resulted in less extreme effects for the top *MIR3681HG* marker, no set of exploratory GWAS consistently eliminated this signal (**Supplementary Materials, Table S4**).

To determine if associations were driven by the imbalance in case:controls, we performed 1:1 and 1:10 matching for each case with population-based common controls via two different commonly used matching methods (Mahalanobis distance and propensity score-based matching). While the Mahalanobis matched cohorts resulted in deflated estimates for the chromosome 2 *MIR3681HG* signal (OR~1.44, 1.46, respectively) closer to the HCV clearance vs. HCV persistence GWAS than all ancestry-matched UKB controls, 1:10 matching resulted in the locus remaining suggestively associated (P=6.76×10^−7^). The use of propensity score-based matching resulted in a further inflation in the effect estimate for the locus (OR~1.76, 1.63, respectively; **Table S4**). It is possible that local ancestry/population structure is driving this signal.

## DISCUSSION

Understanding the epidemiological contexts where common controls risk the internal validity of a GWAS is necessary, especially with the availability of resources like biobanks and national cohorts.^54^ While previous work using population-based controls has suggested non-differential misclassification of outcome can be partially compensated by increased sample sizes and statistical power,^2,6–8,10^ this current work demonstrates that for studies of infectious diseases, the comparison of cases to controls of unknown pathogen exposure can result in spurious associations.

The simulations show that ignoring pathogen exposure among controls can result in a more inflated effect estimate for loci associated with exposure when the prevalence of the pathogen is rare compared to when it is common. Thus, GWAS involving common controls investigating sequelae associated with endemic viruses like cytomegalovirus or Epstein-Barr virus may be less susceptible to exposure-linked selection bias since it is likely all adults have been exposed.^55,56^ However, infectious outcomes specific to HIV, tuberculosis, or malaria need to consider whether the exposure profiles of the selected controls approximate the cases.

Ideally, all controls would be evaluated and tested, but if that is not feasible on a large scale then sampling from high endemic areas or from subpopulations with increased burdens of disease could reduce the risk of spurious associations. Similarly, simulations indicate that epidemiological factors even moderately associated with pathogen exposure in a population where the pathogen of interest is rare can result in spurious genetic associations. This bias may be especially problematic for emerging pathogens like SARS-CoV-2 where certain comorbidities and demographic characteristics may be associated with SARS-CoV-2 exposure (i.e. occupation, living conditions) and the prevalence of the viral exposure depends upon changing political, social, economic, immune and viral factors.^11,57^

For example, in studies of disease severity/death due to COVID-19 involving population based common controls several associations have been observed within loci associated with risk factors for SARS-CoV-2 infection like blood type (e.g., ABO),^6,58,59^ obesity,^60^ alcohol use,^61^ and non-European genetic ancestry.^59,60^ Most notable is the ABO signal in chromosome 9, which is the most significantly associated locus for SARS-CoV-2 infection and significantly associated with both hospitalization and critical illness due to COVID-19 when using population-based controls, according to the COVID-19 Host Genetics Initiative’s meta-analysis results (r6 release).^6^ However, these markers fail to reach nominal statistical significance when controls are limited to non-hospitalized individuals with COVID-19 infection,^6^ suggesting the ABO locus may not be a valid association. Loci associated with risk factors for SARS-CoV-2 could also result in spurious associations for other infectious disease-related outcomes when using common controls, as blood type is similarly associated with susceptibility to various bacterial infections and SARS,^62,63^ obesity with influenza and pneumonia,^64,65^ and alcohol use with contracting tuberculosis,^66^ HIV,^67^ and pneumonia.^68^ However, multiple COVID-19 severity associated loci identified using population-based common controls may reflect real associations, as they were also successfully identified in GWAS limited to SARS-CoV-2 exposed controls (e.g, *LZTFL1, IFNAR2*).

Using empirical GWAS data we show that population-based common controls replicated HCV clearance-associated loci, however markers within the known gene association locus of *IFNL3* were significantly deflated (OR~0.6) as compared to the associations with pathogen exposed controls (OR~0.4), likely due to non-differential misclassification. In contrast, the novel HCV clearance association within *STX18* is likely spurious as it is attenuated after the removal of hemophiliac cases and their matched controls. Relatively rare in the general population, hemophilia is a major risk factor for HCV exposure,^69,70^ and this association may have been driven by enrichment of hemophiliacs, or some other risk factor potentially related to the use of blood products, among cases compared to UKB controls (‘U1~pathogen exposure’). *STX18* is a member of the soluble N-ethylmaleimide-sensitive factor attachment protein receptors (SNARE) protein family and involved in vesicular transport. *STX18* is not known to be associated with HCV clearance or infection and no association was observed for this locus in GWAS involving pathogen exposed controls.

The HCV association with lncRNA *MIR3681HG* remains unclear. The majority of previously published GWAS associations for this locus involve UKB controls,^71–74^ with significantly associated phenotypes including height,^75^ BMI,^76^ and educational attainment.^77^ Systemic differences between the UKB and the UK population have been described, with UKB participants being healthier than the general population (i.e. a ‘healthy volunteer’ bias) and genetic loci identified which are linked participation within the UKB.^74,78–81^ These participation-associated loci have also been linked to educational attainment,^81^ which altogether may explain particularly the biased associations in the *MIR3681HG* locus in our GWAS involving UKB participants given its association with BMI and educational attainment. A reported association with COVID-19 susceptibility is also noted.^82^ Identified in a series of GWAS using cross-sectional snapshots of UKB participant data,^82^ *MIR3681HG* reached genome-wide significance once but failed to remain significant despite increasing cases of COVID-19.^82^

Genetic studies aim to limit spurious findings by setting clear quality control measures, stringent significance thresholds, and requiring replication of findings to reduce the costly consequences of implementing translational analysis of spurious signals. These findings do not detract from the utility of GWAS involving population-based common controls, which remains an important and valuable approach to interrogate the genetic risk factors of human health and disease. Rather, we recommend that findings from infectious disease-focused association studies involving common controls be interpreted with context and caution.

In sum, the use of population-based common controls may be more problematic for infectious disease-focused GWAS than previously described depending on the probability of exposure to the infectious pathogen.^7,8^ While true disease-associated loci can be identified, concerns related to selection bias, confounding, and misclassification are exacerbated by the inability to account for pathogen exposure among common controls.^11^ Efforts to increase the selection of controls from areas with high prevalence or endemicity of disease or selection of older age participants if there is known childhood exposure may ameliorate the risk of false findings. Otherwise, controls should be carefully selected and screened for pathogen exposure.

## Data Availability

Access to individual-level phenotypic and genetic data from HCV extended genetics consortium individuals can be requested via dbGaP (phs000248.v1.p1). Access to individual-level data from the UKB can be requested at https://www.ukbiobank.ac.uk. Summary statistics for each GWAS performed can be viewed and downloaded at https://my.locuszoom.org/ (Study name: HCV Consortium vs. UKB). Code used to perform simulation experiments, bipartite PCA-based matching, and gnomAD MAF-based variant-level filtering can be accessed at https://github.com/dduchen/Population_Based_Controls_GWAS_ID_Manuscript.

## Declaration of interests

All authors declare no competing interests.

## Acknowledgments

2R01AI148049 along with a COVID-19 supplement under the same grant number (D.L.T, P.D., G.L.W) and Burroughs-Wellcome Fund, MD-GEM training grant (D.D.). Access and use of the UK Biobank was approved using application number 17712. G.L.W. was additionally supported by the National Human Genome Research Institute (NHGRI) grant R35HG011944.

## Author contributions

The study was designed by D.D., C.C., G.L.W., and P.D. Data collection was led by (D.L.T., C.L.T, N.C., P.K., P.D.). Simulation was performed by D.D. Statistical analyses were designed and performed by D.D., C.C., G.L.W., and P.D. Manuscript was first drafted by D.D., C.C., G.L.W., and P.D. All authors contributed to the final manuscript.

## Notes

### Competing Interest Statement

The authors have declared no competing interest.

### Funding Statement

Funding for this study:
2R01AI148049 along with a COVID-19 supplement under the same grant number (D.L.T, P.D., G.L.W) and Burroughs-Wellcome Fund, MD-GEM training grant (D.D.). G.L.W. was additionally supported by the National Human Genome Research Institute (NHGRI) grant R35HG011944.

### Author Declarations

Access and use of data from the UK Biobank was approved using application number 17712. Consent was obtained for genotyping as approved by the governing IRB and DNA provided to Johns Hopkins School of Medicine without identifiers, as described previously (doi: 10.1053/j.gastro.2018.12.014, 10.7326/0003-4819-158-4-201302190-00003).

