## Supplementary Figures/Tables for "Pathogen exposure misclassification can bias association signals in GWAS of infectious diseases when using population-based common controls"

**Supplemental Materials:**

**Legend of Figures**

- **Figure S1:** Principal components analysis plot of the first two principal components estimated from 1,739 HCV Consortium individuals and 370,702 UKB individuals.
- **Figure S2**: Scree plot detailing the variance explained by the first twenty principal components estimated from 1,739 HCV Consortium individuals and 370,702 UKB individuals.
- **Figure S3**: Manhattan plot of the GWAS comparing HCV clearance cases to UKB controls sharing <2% IBD.
- **Figure S4**: Manhattan plot of GWAS comparing HCV clearance cases to stringent QC-passed UKB controls.
- **Figure S5:** Manhattan plot of the GWAS comparing HCV clearance cases to propensity score matched (1:10) UKB controls.
- **Figure S6:** Manhattan plot of the GWAS comparing HCV clearance cases to propensity score matched (1:1) UKB controls.
- **Figure S7:** Manhattan plot of the GWAS comparing HCV clearance cases to Mahalanobis distance matched (1:1) UKB controls.
- **Figure S8:** Manhattan plot of the GWAS comparing HCV clearance cases to Mahalanobis distance matched (1:10) UKB controls.
- **Figure S9:** Manhattan plot of the GWAS comparing HCV clearance cases without hemophilia to propensity score matched (1:10) UKB controls.
- **Figure S10:** Manhattan plot of the GWAS comparing all HCV Consortium cohort individuals to UKB Controls.
- **Figure S11:** Manhattan plot of the GWAS comparing HCV clearance cases to ancestry matched UKB controls highlighting markers significantly heterogenous compared to the HCV clearance vs. HCV persistence GWAS.
- **Figure S12:** Distribution of observed odds ratios for a non-outcome associated locus moderately associated with pathogen exposure across varying pathogen exposure prevalence.
- **Figure S13:** Distribution of observed odds ratios for a non-outcome associated locus strongly associated with pathogen exposure across a variable number of cases.
- **Figure S14:** Quantile-quantile plots of main GWAS performed.
- **Figure S15:** Distribution of observed odds ratios for a non-outcome associated locus moderately associated with pathogen exposure across varying post-exposure outcome prevalence across the cohort.
- **Figure S16:** Distribution of observed odds ratios for a non-outcome associated locus strongly associated with pathogen exposure across varying post-exposure outcome prevalence across the cohort.
- **Figure S17:** Variability of effect estimates for HCV clearance-associated loci of interest using alternative matched case-control cohorts.

**Legend of Tables**

- **Table S1:** Heterogeneity estimates across simulated cohorts.
- **Table S2:** Suggestively associated markers (P<5x10^-5^) from the GWAS comparing HCV clearance cases to ancestry-matched UKB controls. *[separate file/link].*
- **Table S3:** Variability in effect estimates of known loci across performed GWAS.
- **Table S4:** Variability in effect estimates of novel loci across performed GWAS.
- **Table S5:** Additional simulation scenario (Scenario 3).

**Figures**

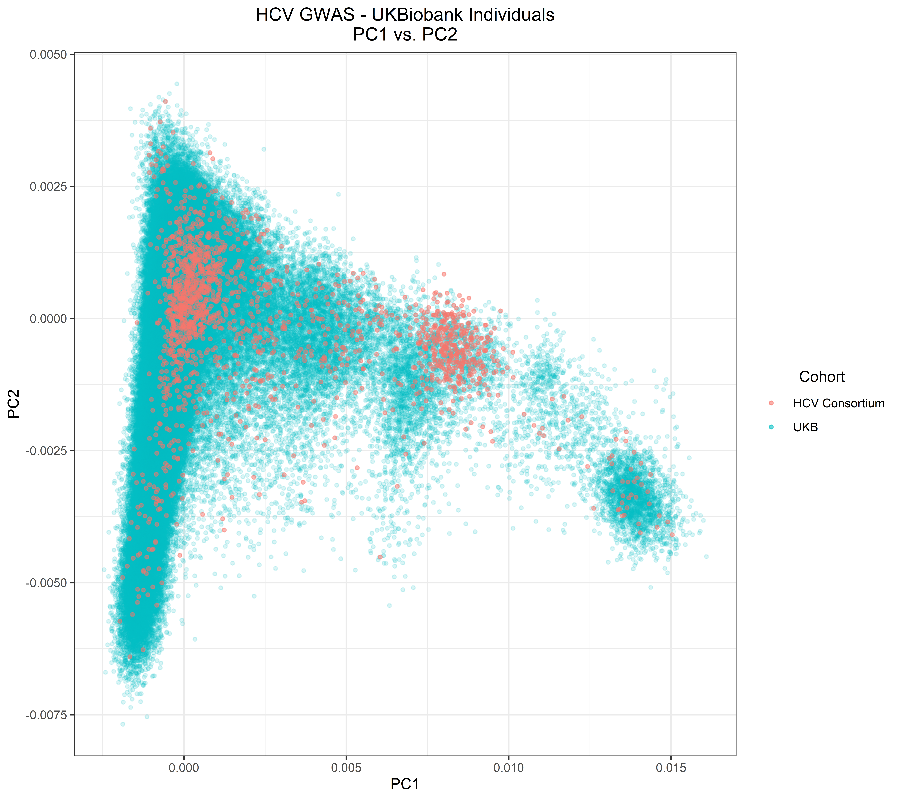

**Figure S1**: Principal components analysis plot of the first two principal components estimated from 1,739 HCV Consortium individuals and 370,702 UKB individuals. The X axis reflects the estimated eigenvalues of the first principal component. The Y axis reflects the estimated eigenvalues from the second principal component. Points reflect individuals from the HCV consortium (red) or UK Biobank (blue).

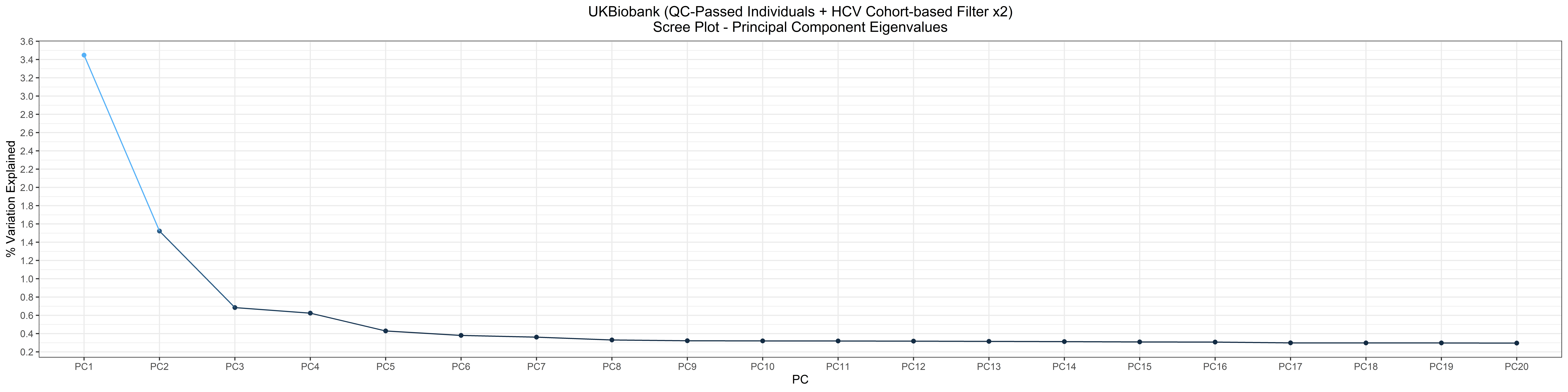

**Figure S2**: Scree plot detailing the variance explained by the first twenty principal components estimated from 1,739 HCV Consortium individuals and 370,702 UKB individuals. The X axis reflects each principal component. The Y axis reflects the percentage of the total variance explained by each derived principal component.

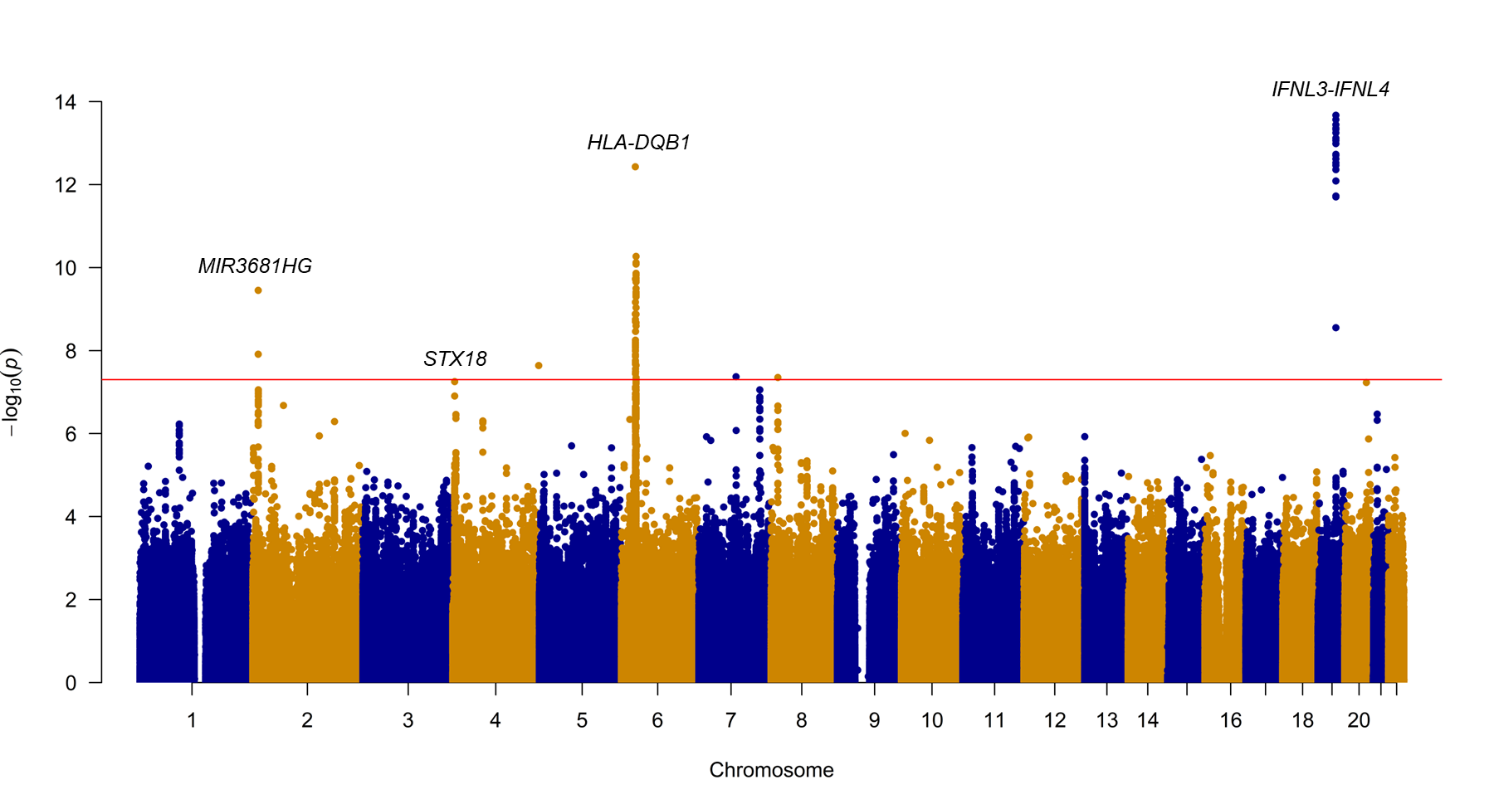

**Figure S3**: Manhattan plot of the GWAS comparing HCV clearance cases to UKB controls sharing <2% IBD. This GWAS compared HCV clearance cases (N=702) to propensity score-based matched UKB controls further limited to UKB controls sharing IBD proportions under 2% (N=6,512). Each point reflects the P value for a marker from across the genome, ordered along the X axis by chromosomal position. The Y axis reflects the -log(P value), with the most significantly associated markers rising above the genome-wide significance threshold (P<5x10^-8^) indicated by the red line.
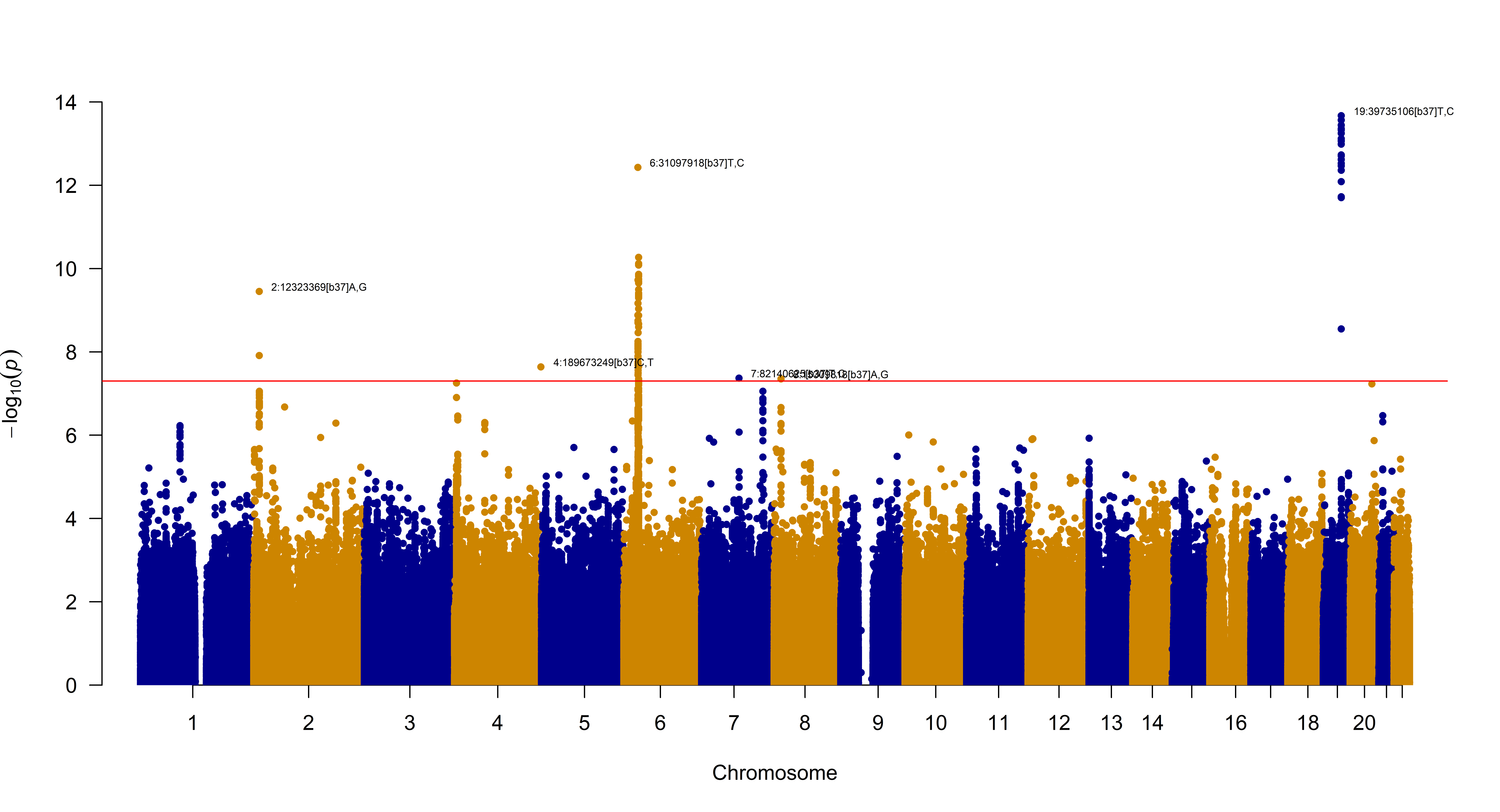

**Figure S4**: Manhattan plot of GWAS comparing HCV clearance cases to stringent QC-passed UKB controls. This GWAS compared HCV clearance individuals (N=702) to the set of UKB individuals who passed stringent QC metrics (N=364,308). Each point reflects the P value for a marker from across the genome, ordered along the X axis by chromosomal position. The Y axis reflects the -log(P value), with the most significantly associated markers rising above the genome-wide significance threshold (P<5x10^-8^) indicated by the red line.

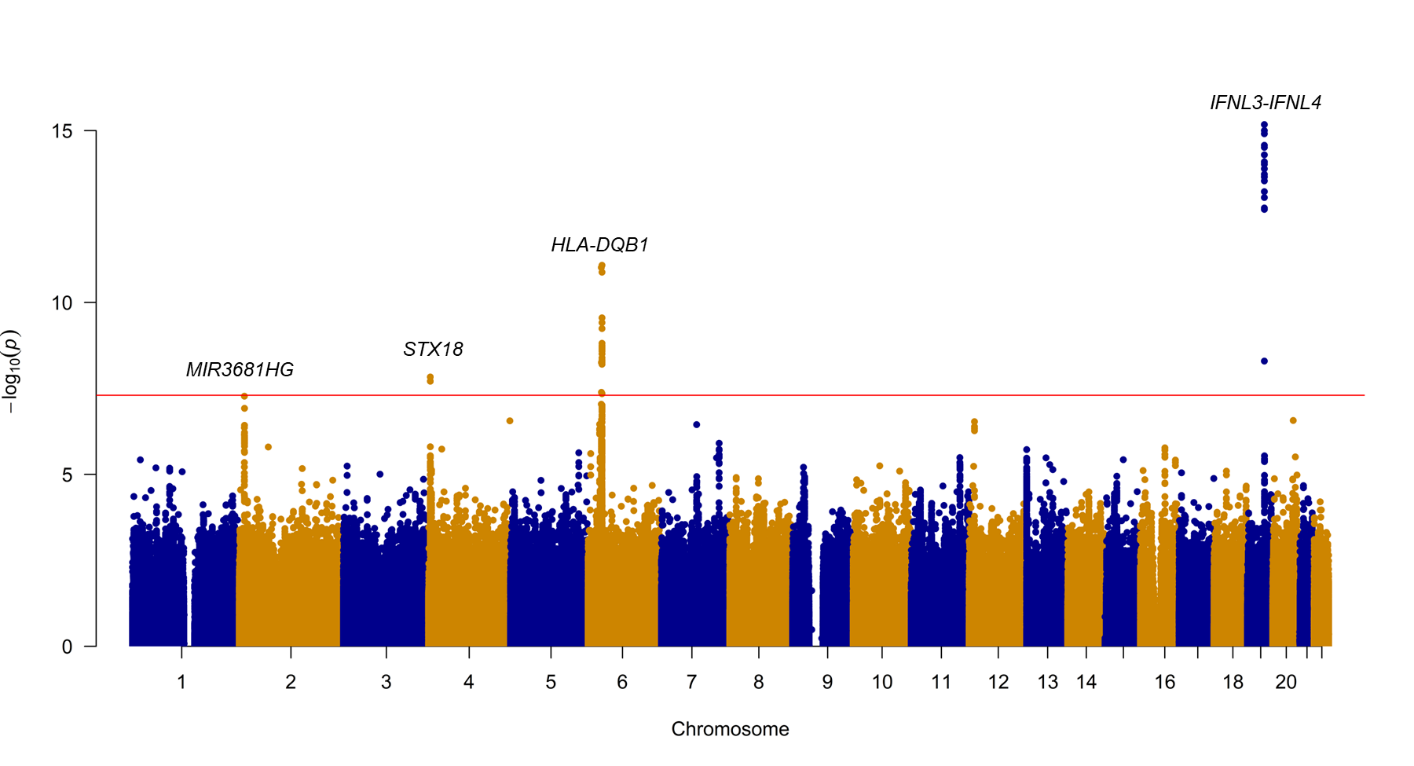

**Figure S5:** Manhattan plot of the GWAS comparing HCV clearance cases to propensity score matched (1:10) UKB controls. This GWAS compared HCV clearance cases (N=702) to propensity score-based matched UKB controls at a ratio of 1:10 (N=6,666). Each point reflects the P value for a marker from across the genome, ordered along the X axis by chromosomal position. The Y axis reflects the -log(P value), with the most significantly associated markers rising above the genome-wide significance threshold (P<5x10^-8^) indicated by the red line.

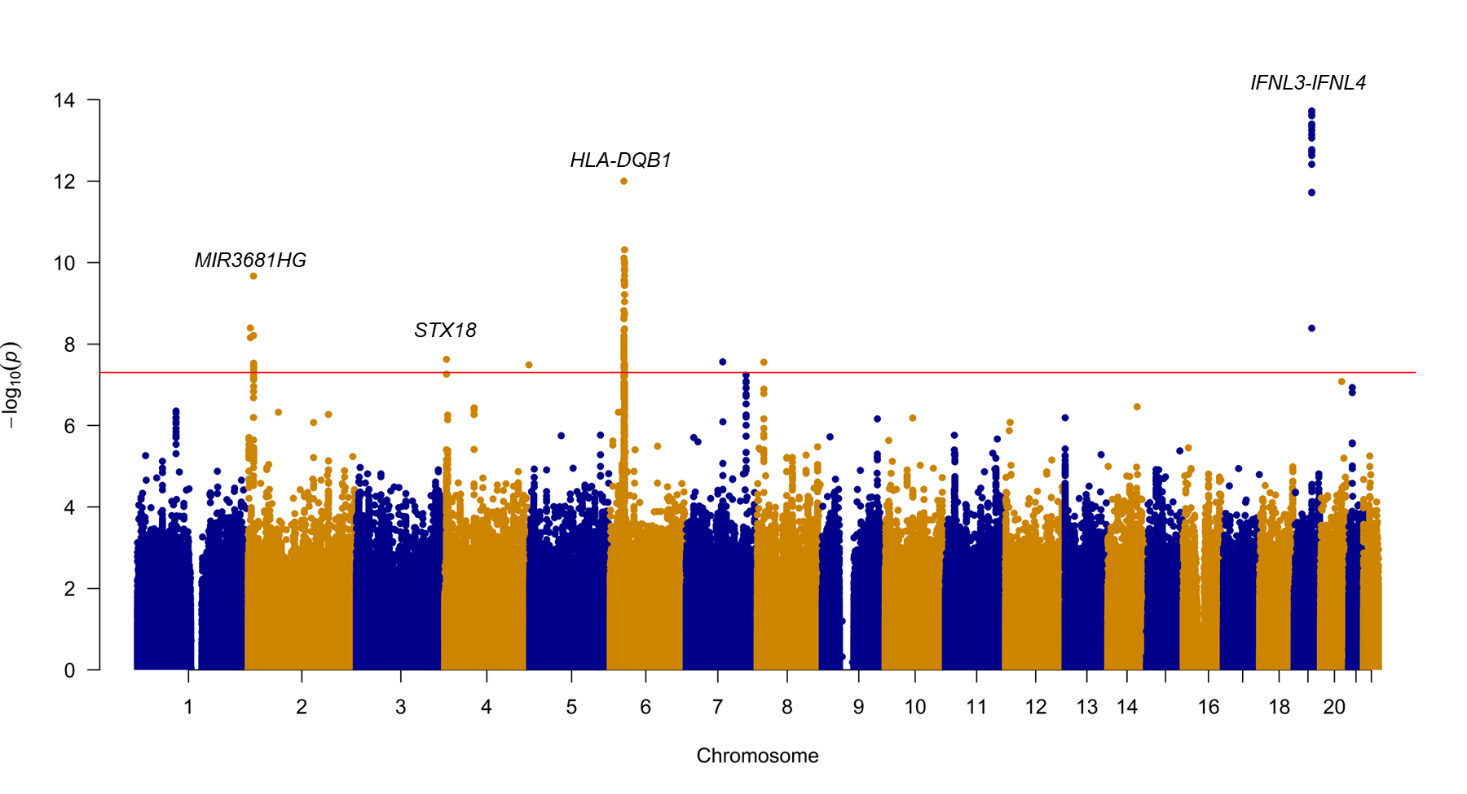

**Figure S6:** Manhattan plot of the GWAS comparing HCV clearance cases to propensity score matched (1:1) UKB controls. This GWAS compared HCV clearance cases (N=702) to propensity score matched UKB controls at a ratio of 1:1 (N=702). Each point reflects the P value for a marker from across the genome, ordered along the X axis by chromosomal position. The Y axis reflects the -log(P value), with the most significantly associated markers rising above the genome-wide significance threshold (P<5x10^-8^) indicated by the red line.

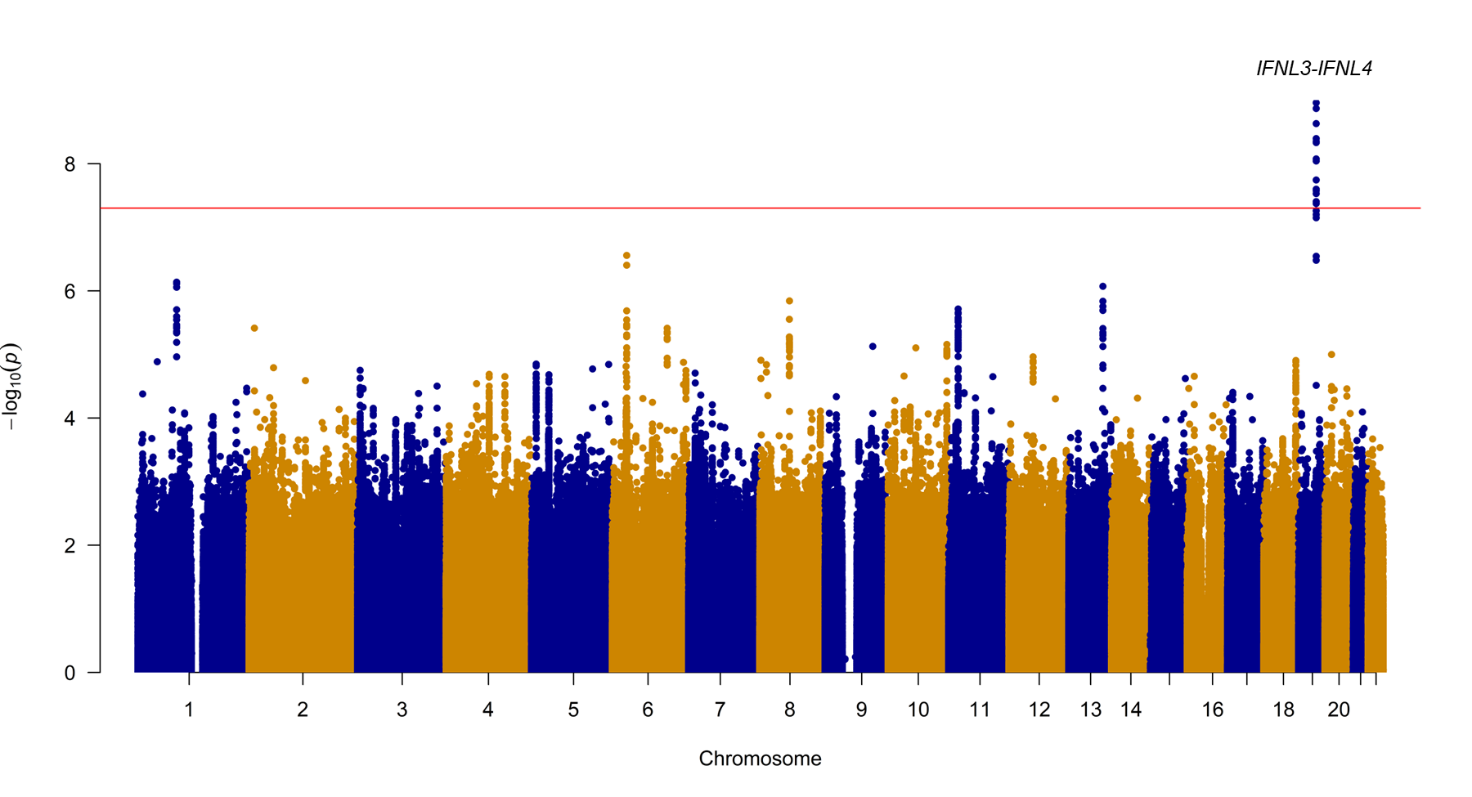

**Figure S7:** Manhattan plot of the GWAS comparing HCV clearance cases to Mahalanobis distance matched (1:1) UKB controls. This GWAS compared HCV clearance cases (N=702) to Mahalanobis distance-based matched UKB controls at a ratio of 1:1 (N=702). Each point reflects the P value for a marker from across the genome, ordered along the X axis by chromosomal position. The Y axis reflects the -log(P value), with the most significantly associated markers rising above the genome-wide significance threshold (P<5x10^-8^) indicated by the red line.

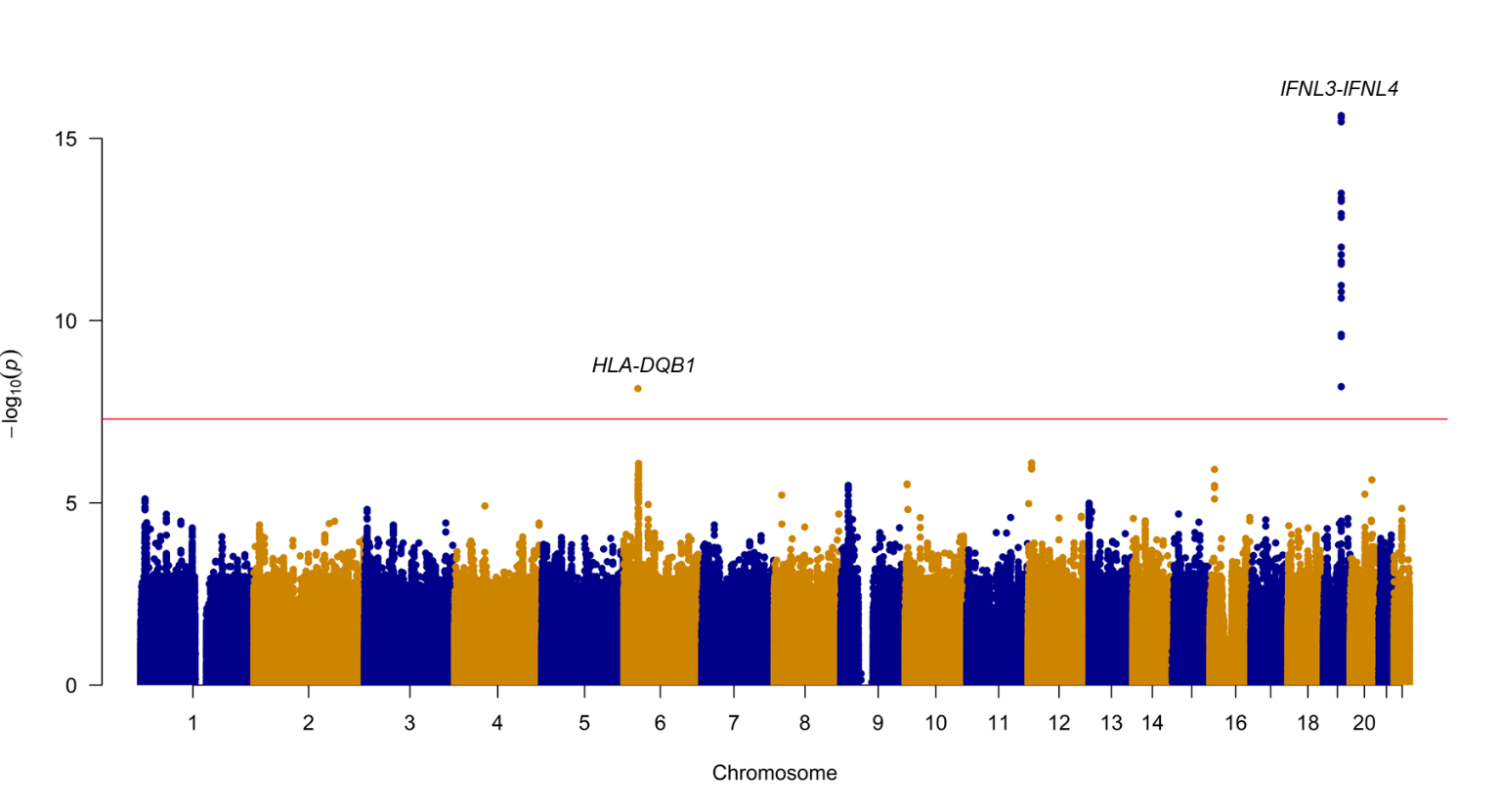

**Figure S8**: Manhattan plot of the GWAS comparing HCV clearance cases to Mahalanobis distance matched (1:10) UKB controls. This GWAS compared HCV clearance cases (N=702) to Mahalanobis distance-based matched UKB controls at a ratio of 1:10 (N=6,633). Each point reflects the P value for a marker from across the genome, ordered along the X axis by chromosomal position. The Y axis reflects the -log(P value), with the most significantly associated markers rising above the genome-wide significance threshold (P<5x10^-8^) indicated by the red line.

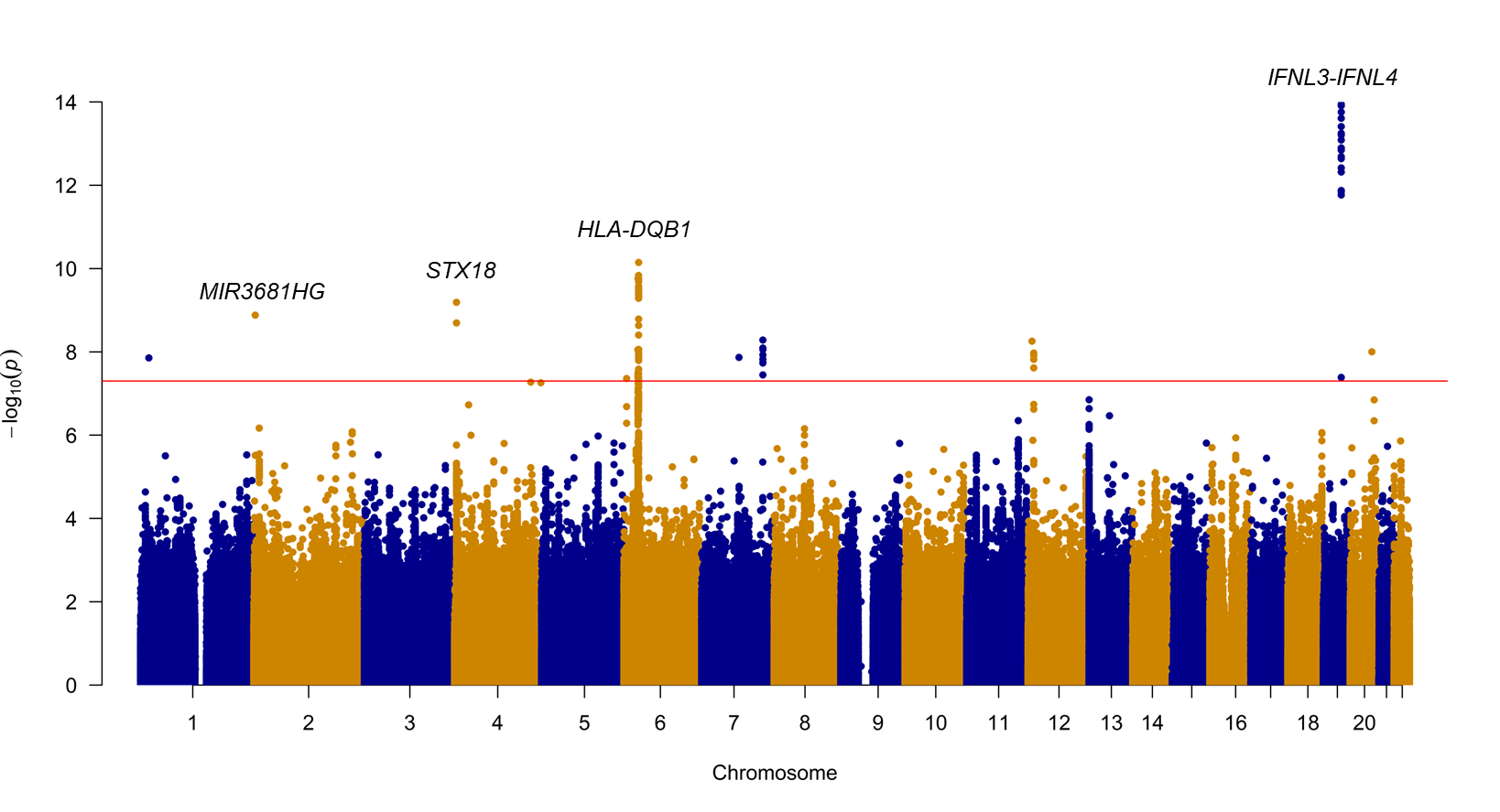

**Figure S9**: Manhattan plot of the GWAS comparing HCV clearance cases without hemophilia to propensity score matched (1:10) UKB controls. This GWAS compared HCV clearance cases recruited from non-hemophilia cohorts (N=513) to propensity score-based matched UKB controls at a ratio of 1:10 (N=4,908). Each point reflects the P value for a marker from across the genome, ordered along the X axis by chromosomal position. The Y axis reflects the -log(P value), with the most significantly associated markers rising above the genome-wide significance threshold (P<5x10^-8^) indicated by the red line.

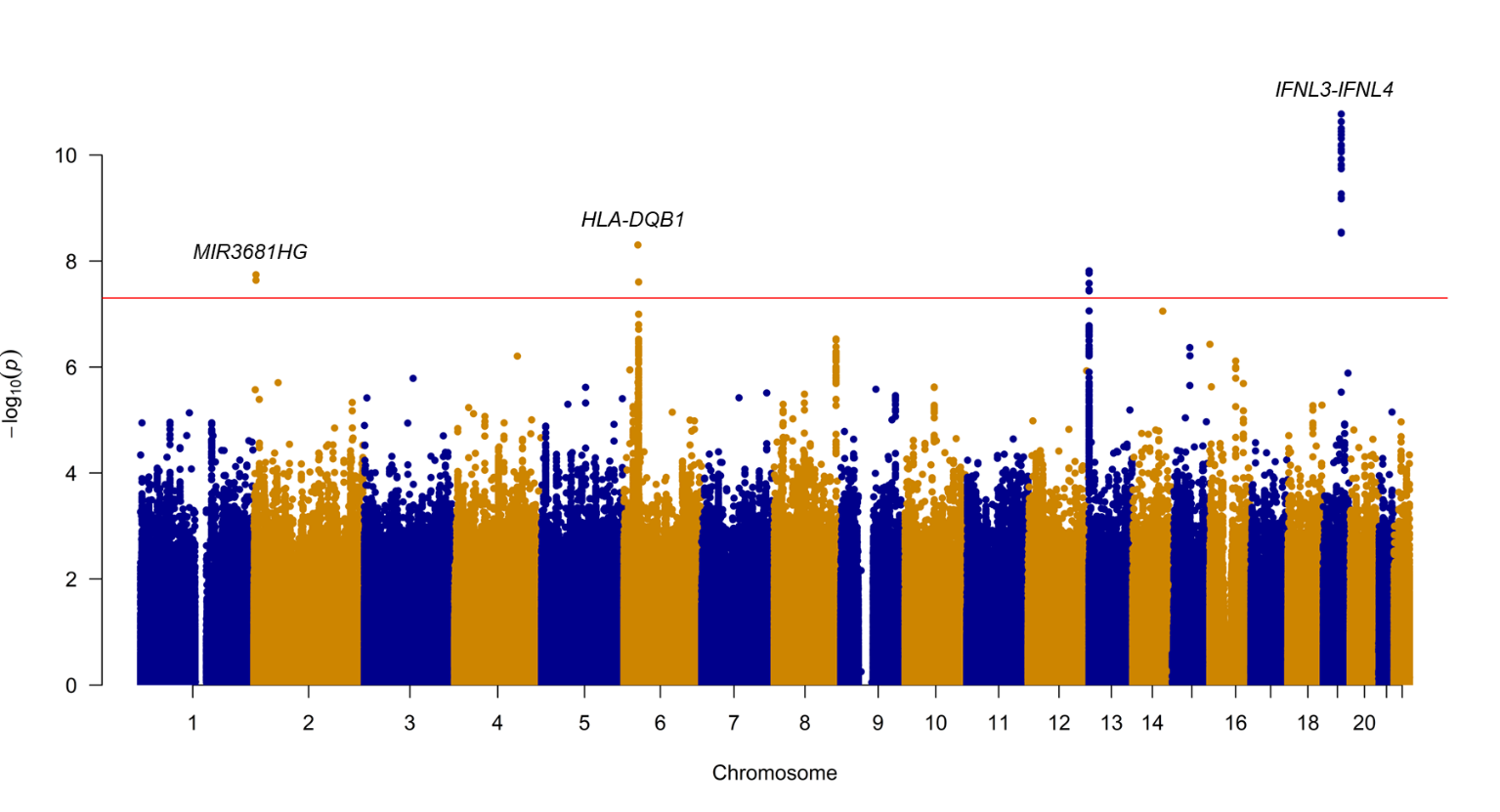

**Figure S10**: Manhattan plot of the GWAS comparing all HCV Consortium cohort individuals to UKB Controls. This GWAS compared HCV Consortium individuals (N=1,739) to the ancestry-matched UKB controls (N=370,702). Each point reflects the P value for a marker from across the genome, ordered along the X axis by chromosomal position. The Y axis reflects the -log(P value), with the most significantly associated markers rising above the genome-wide significance threshold (P<5x10^-8^) indicated by the red line.

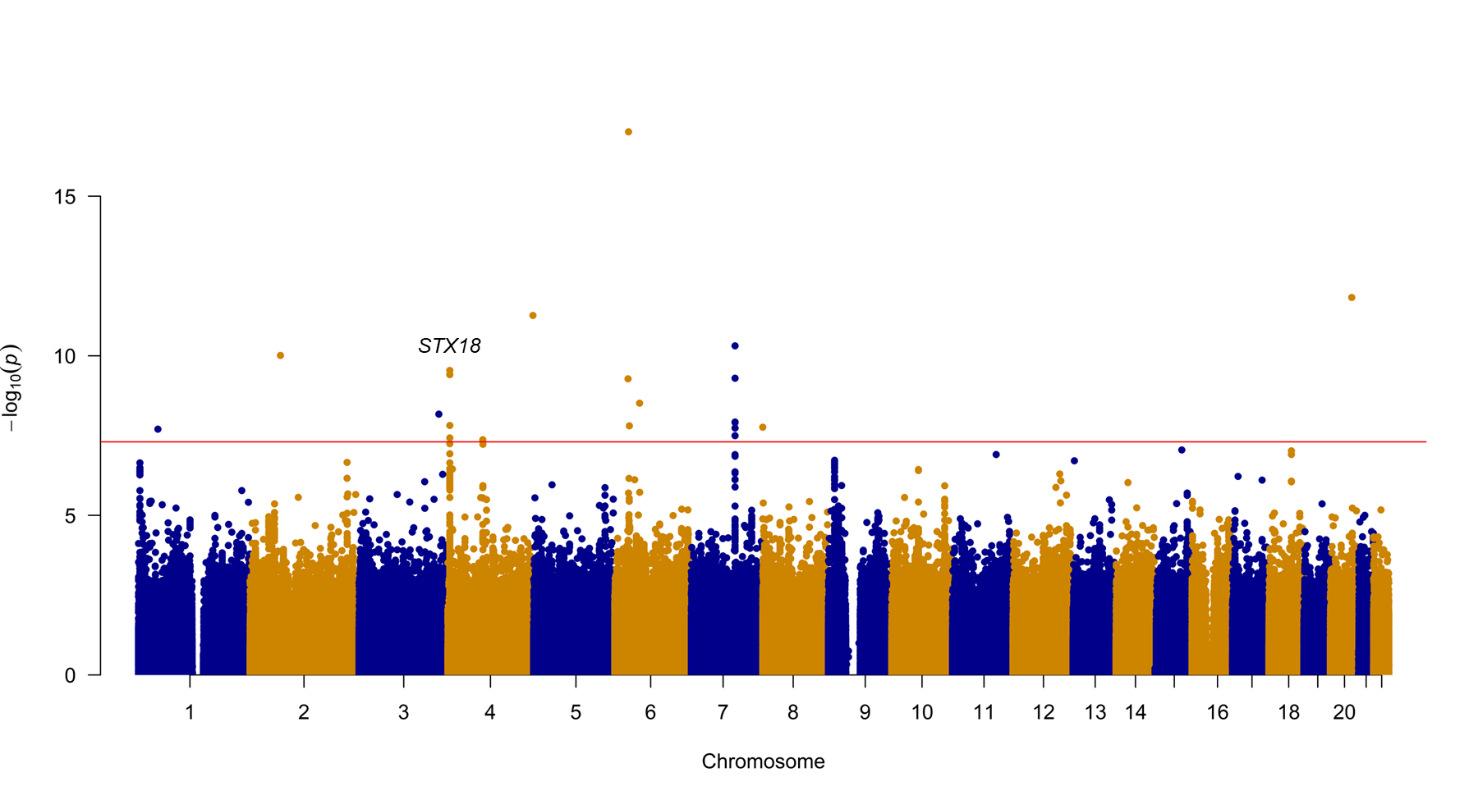

**Figure S11**: Manhattan plot of the GWAS comparing HCV clearance cases to ancestry matched UKB controls highlighting markers significantly heterogenous compared to the HCV clearance vs. HCV persistence GWAS. This GWAS compared HCV clearance individuals (N=702) to the ancestry-matched UKB controls (N=370,702). Each point reflects the P value for a marker from across the genome, ordered along the X axis by chromosomal position. Green points reflect markers with significant Cochrane’s Q heterogeneity estimates (P<0.05). The Y axis reflects the -log (P value), with the most significantly associated markers farthest away from 0 and outside the genome-wide significance threshold (P<5x10-8), indicated by the red lines. HCV clearance associated loci of interest have been highlighted and annotated with their nearest/overlapping gene name, with the novel HCV clearance-associated loci highlighted in the bottom panel.

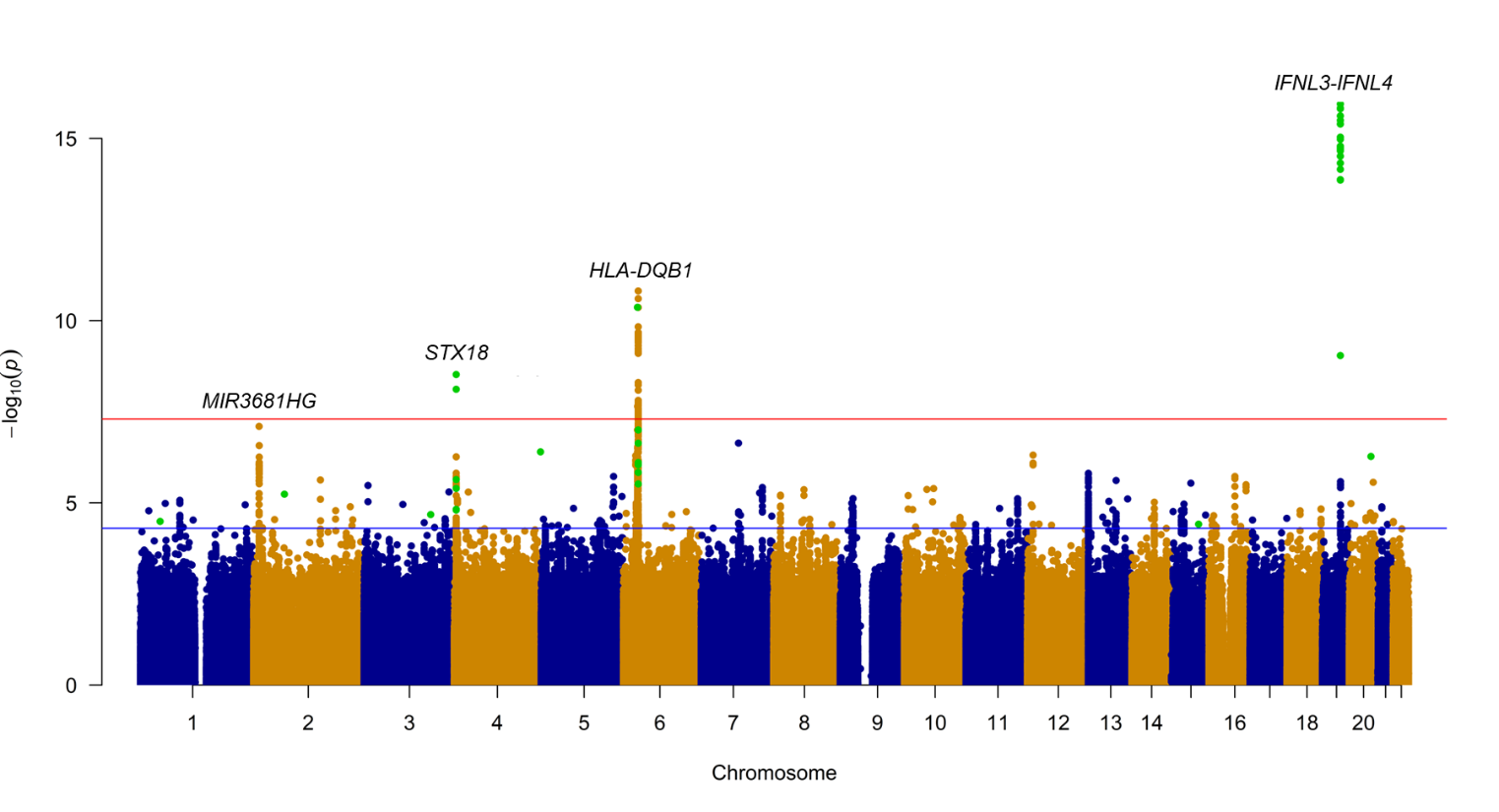

**Figure S12**: Distribution of observed odds ratios for a non-outcome associated locus moderately associated with pathogen exposure across varying pathogen exposure prevalence. Comparisons involved 20,000 cases and equal numbers of well-characterized or population-based controls (left) or 20,000 cases and equal numbers of well-characterized and 200,000 population-based controls (right), assuming a moderate U1~’Pathogen exposure’ relationship. Boxplots reflect distribution of odds ratios comparing cases to population-based (red) or well-characterized controls (yellow). Reported P-values derived from a Z score based on the difference between averaged beta estimates when using population-based controls vs. well-characterized controls.

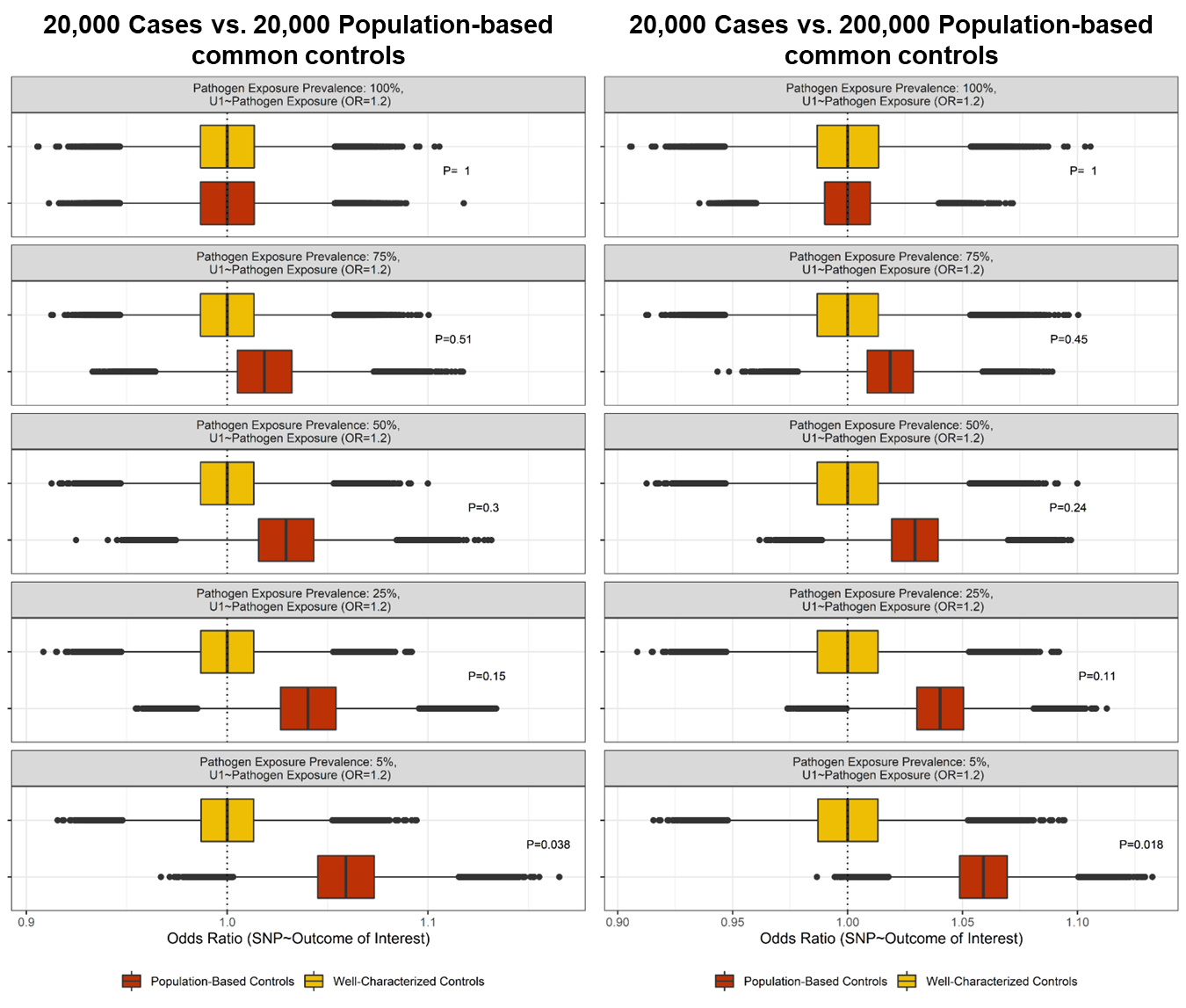

**Figure S13**: Distribution of observed odds ratios for a non-outcome associated locus strongly associated with pathogen exposure across a variable number of cases. The title of each panel details the number of cases compared to population-based or well-characterized controls. Boxplots reflect odds ratios obtained when comparing the relevant number of cases to 20,000 population-based (red) or 20,000 well-characterized controls (yellow). Reported P-values derived from a Z score based on the difference between averaged beta estimates when using population-based controls vs. well-characterized controls.

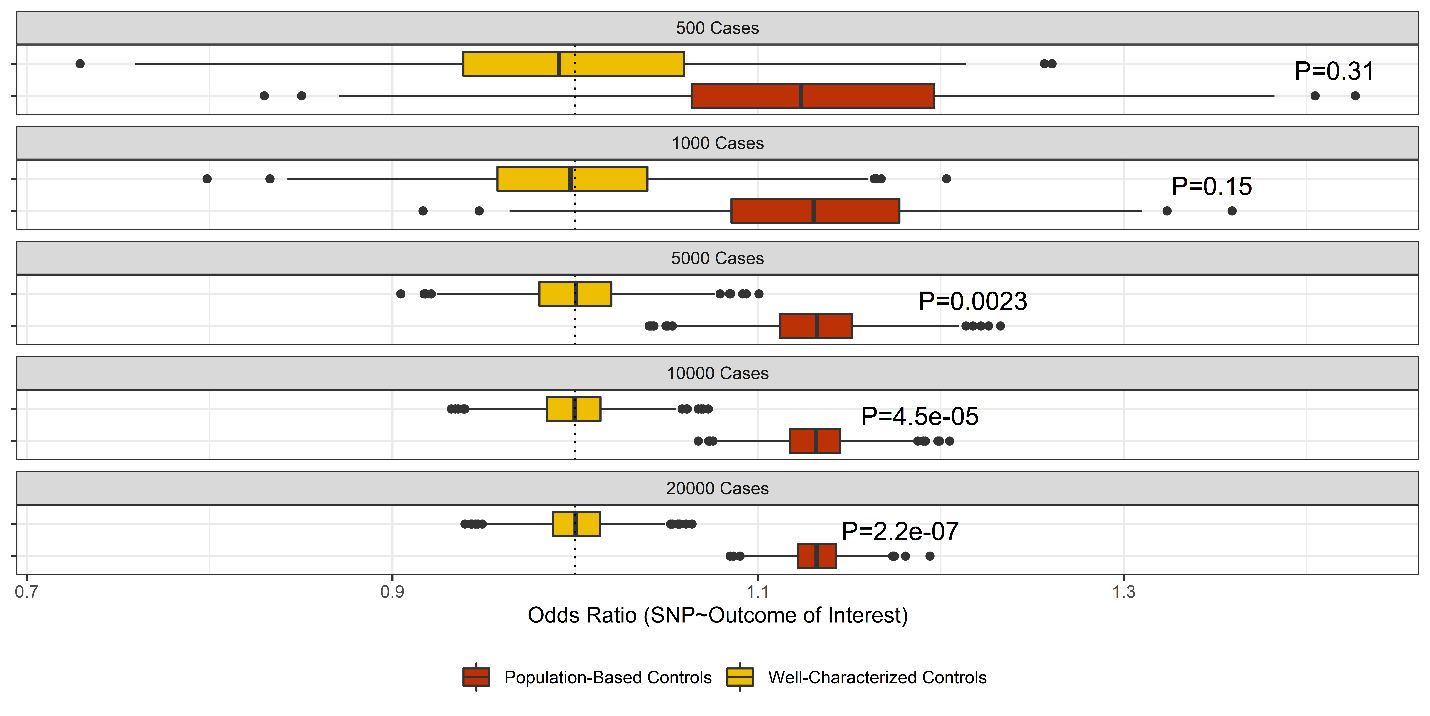

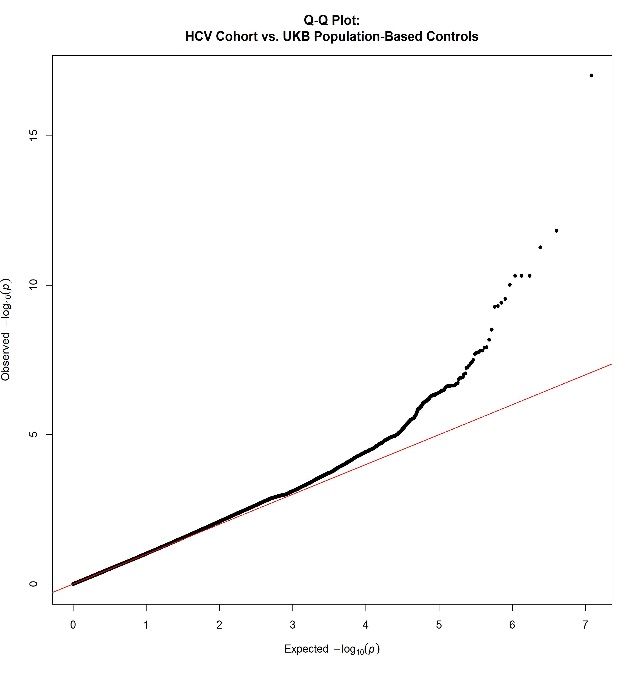

HCV Consortium vs. UKB controls (λ=1.013)

HCV clearance vs. HCV persistence (λ=1.013)

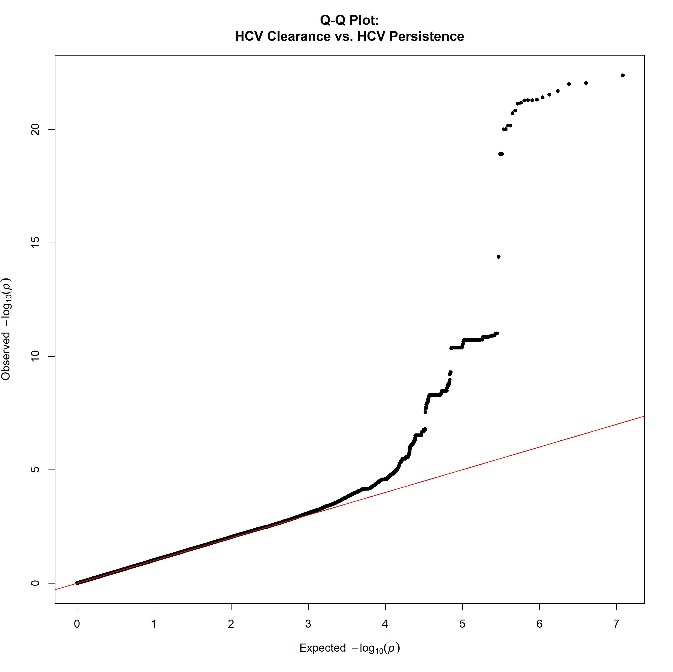

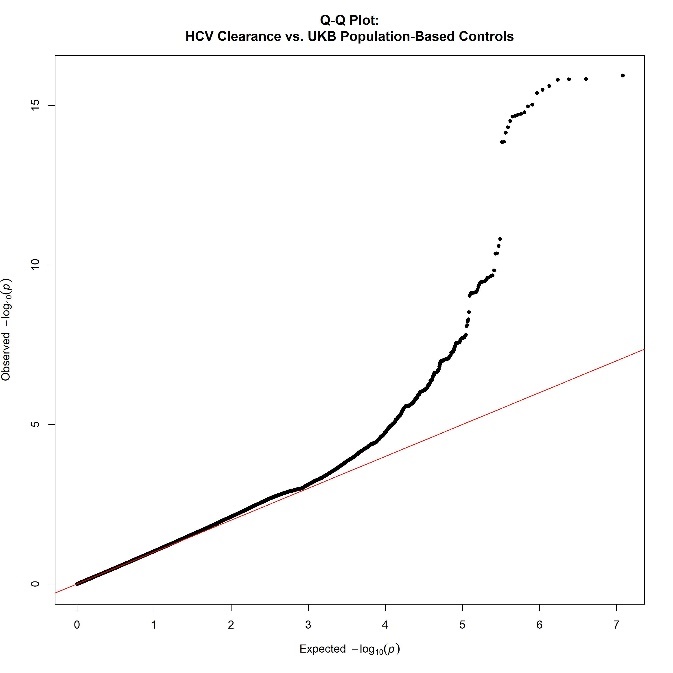

HCV clearance vs. UKB controls (λ=1.026)

**Figure S14**: Quantile-quantile plots of main GWAS performed. The X axis reflects the predicted and Y axis the observed p-values associated with variants across the genome from the original HCV Clearance (N=702) vs. HCV persistence (N=1,037) GWAS (top left), the HCV clearance vs. ancestry matched population based UKB controls (N=370,702) GWAS (top right), the full HCV consortium (N=1,739) vs. the ancestry matched population based UKB controls GWAS (bottom).

**Figure S15**. Distribution of observed odds ratios for a non-outcome associated locus moderately associated with pathogen exposure across varying post-exposure outcome prevalence across the cohort. Comparisons involved 20,000 cases and equal numbers of well-characterized or population-based controls (left) or equal numbers of well-characterized and 200,000 population-based controls (right) assuming a moderate U1~’Pathogen exposure’ relationship. Boxplots reflect distribution of odds ratios comparing cases to population-based (red) or well-characterized controls (yellow). Reported P-values derived from a Z score based on the difference between averaged beta estimates when using population-based controls vs. well-characterized controls.

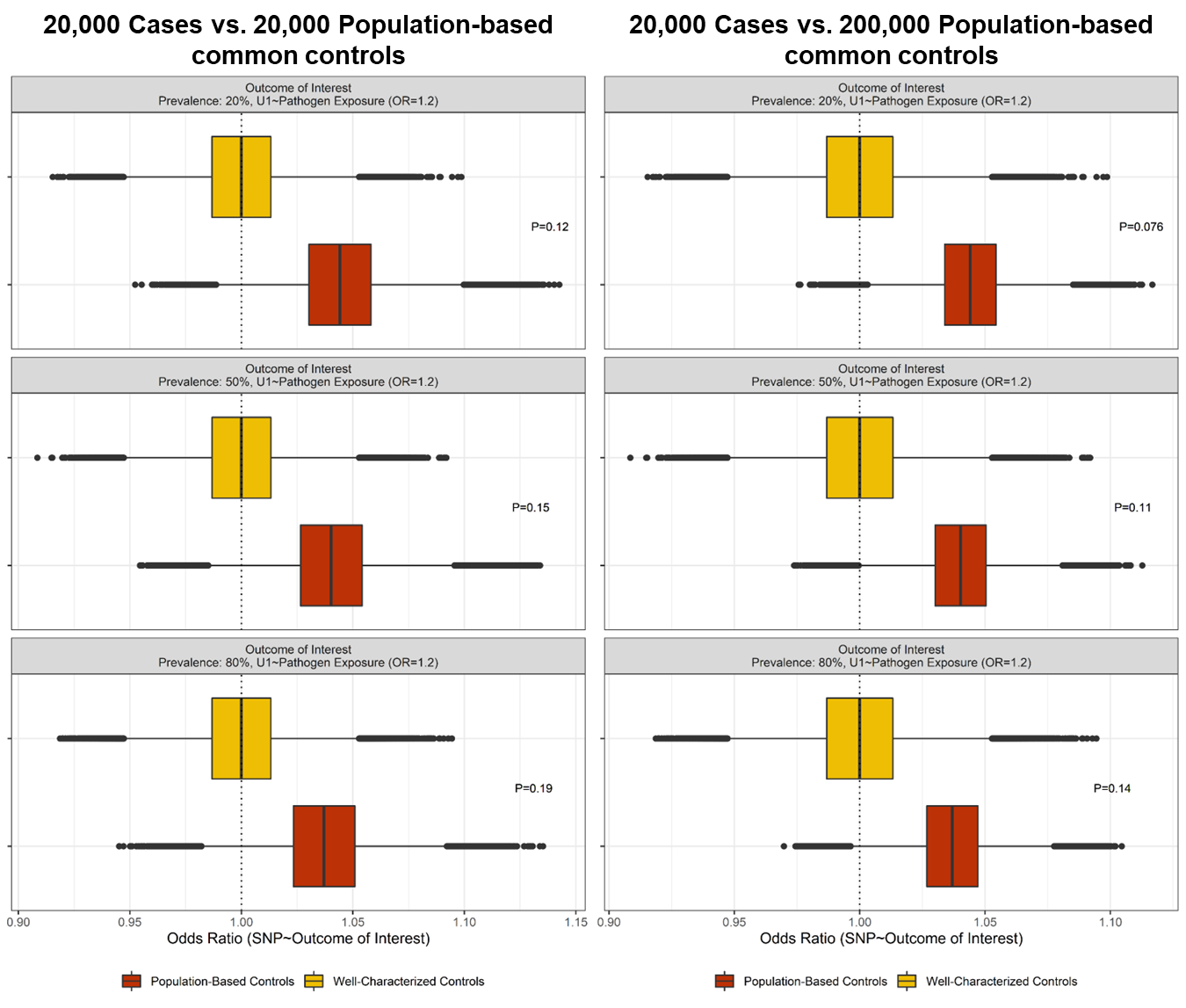

**Figure S16**. Distribution of observed odds ratios for a non-outcome associated locus strongly associated with pathogen exposure across varying post-exposure outcome prevalence across the cohort. Comparisons involved 20,000 cases and equal numbers of well-characterized or population-based controls (left) or 20,000 cases and equal numbers of well-characterized and 200,000 population-based controls (right) assuming a strong U1~’Pathogen exposure’ relationship. Boxplots reflect distribution of odds ratios comparing cases to population-based (red) or well-characterized controls (yellow). Reported P-values derived from a Z score based on the difference between averaged beta estimates when using population-based controls vs. well-characterized controls.

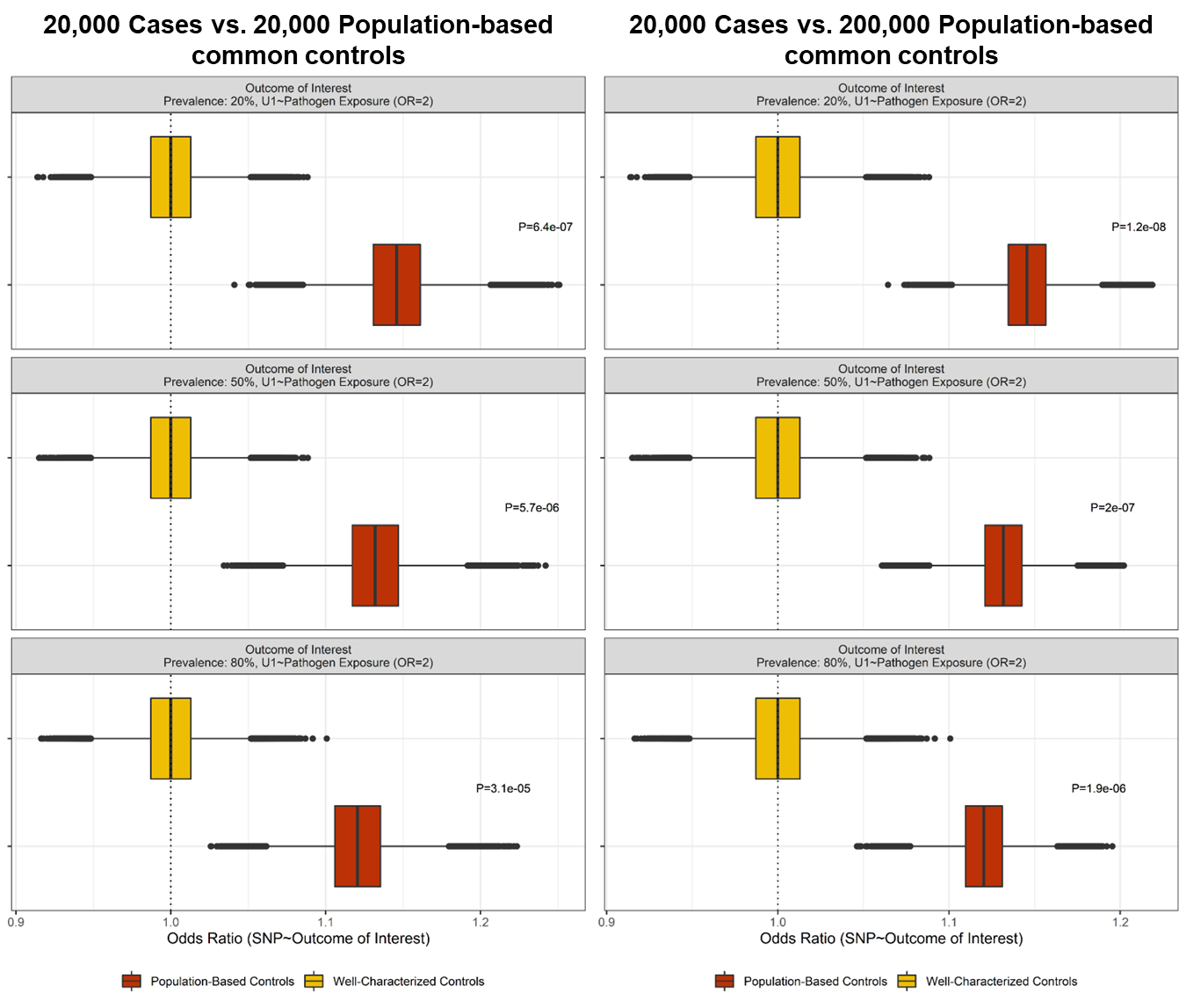

**Figure S17**: Variability of effect estimates for HCV clearance-associated loci of interest using alternative matched case-control cohorts. Distribution of all observed beta estimates (log OR) for the top HCV clearance associated loci (left panel) and, separately by being genome-wide significant (red) or suggestive (orange) (right panel). Distribution of effect estimates broken down by matching method (Mahalanobis distance, propensity score) are shown for both the left and right series of boxplots. In the left panel, the GWAS-derived beta estimates from the analysis of HCV clearance cases vs. persistently infected controls is shown as a vertical light blue line and the beta estimates from the comparison of HCV clearance cases vs. ancestry-matched UKB population-based UKB controls is shown as a vertical red line. CoV: coefficient of variation.

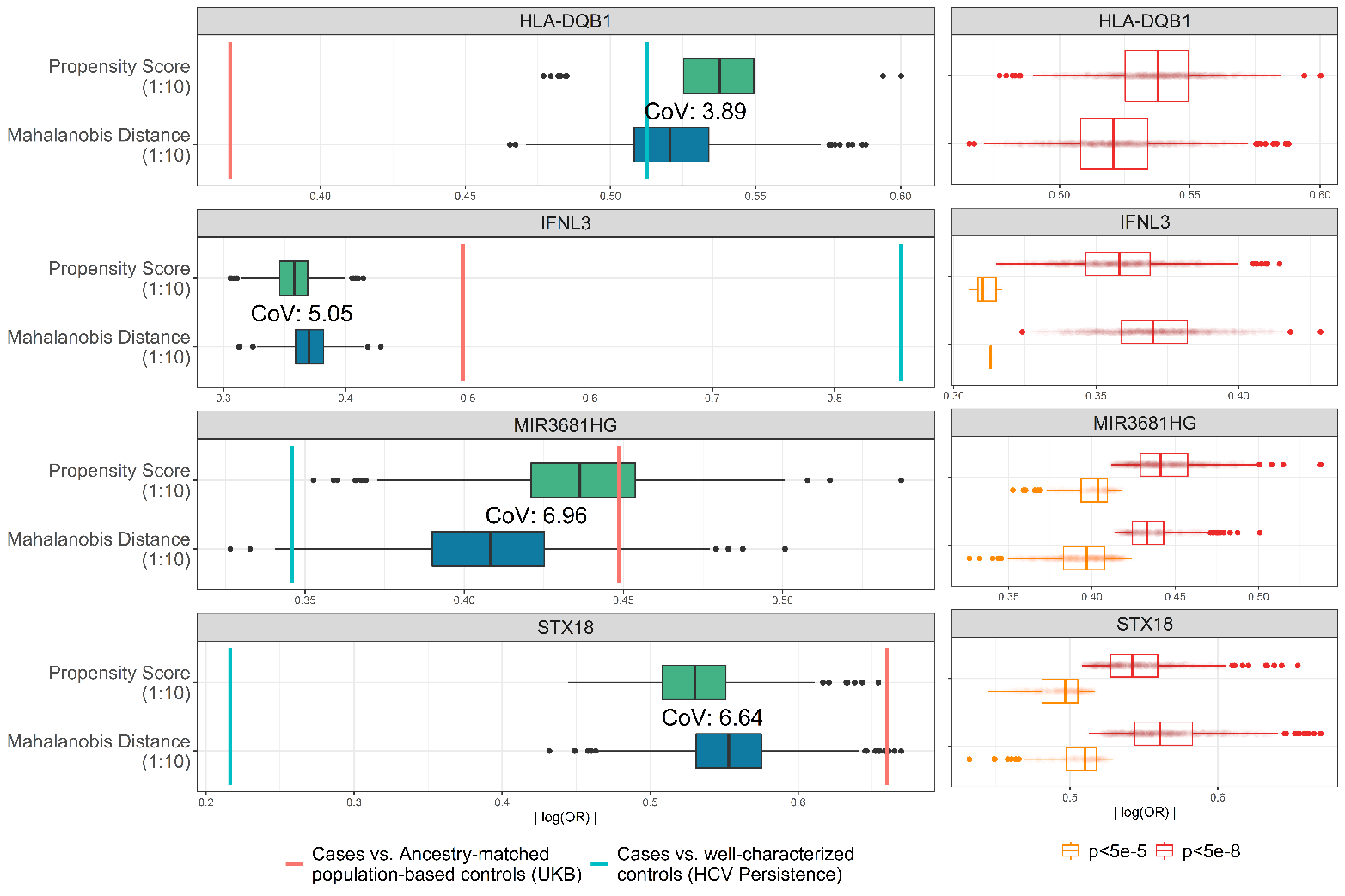

| **Scenario** | **Controls** | **Parameter Value** | **U1~Pathogen Exposure** | **OR** | **Tau^2^** | **I^2^** |
| --- | --- | --- | --- | --- | --- | --- |
| 1 | Pathogen exposed | U1~Pathogen Exposure | OR=1 | 1.00 | 0 | 0.0% |
|  |  |  | OR=1.2 | 1.00 | <0.001 | 4.8% |
|  |  |  | OR=2 | 1.00 | <0.001 | 1.0% |
|  | Population-based | U1~Pathogen Exposure | OR=1 | 1.00 | 0 | 0.0% |
|  |  |  | OR=1.2 | 1.04 | <0.001 | 4.6% |
|  |  |  | OR=2 | 1.13 | <0.001 | 1.8% |
| 2 | Pathogen exposed | Pathogen Exposure Prevalence: 5% | OR=1.2 | 1.00 | 0 | 0.0% |
|  | Population-based |  |  | 1.06 | 0 | 0.0% |
|  | Pathogen exposed |  | OR=2 | 1.00 | 0 | 0.0% |
|  | Population-based |  |  | 1.20 | <0.001 | 6.9% |
|  | Pathogen exposed | Pathogen Exposure Prevalence: 25% | OR=1.2 | 0.00 | 0 | 0.0% |
|  | Population-based |  |  | 1.04 | <0.001 | 1.2% |
|  | Pathogen exposed |  | OR=2 | 1.00 | <0.001 | 6.8% |
|  | Population-based |  |  | 1.13 | <0.001 | 1.5% |
|  | Pathogen exposed | Pathogen Exposure Prevalence: 50% | OR=1.2 | 1.00 | 0 | 0.0% |
|  | Population-based |  |  | 1.03 | <0.001 | 2.9% |
|  | Pathogen exposed |  | OR=2 | 1.00 | <0.001 | 6.1% |
|  | Population-based |  |  | 1.09 | <0.001 | 3.8% |
|  | Pathogen exposed | Pathogen Exposure Prevalence: 75% | OR=1.2 | 1.00 | <0.001 | 9.4% |
|  | Population-based |  |  | 1.02 | 0 | 0.0% |
|  | Pathogen exposed |  | OR=2 | 1.00 | 0 | 0.0% |
|  | Population-based |  |  | 1.06 | 0 | 0.0% |
|  | Pathogen exposed | Pathogen Exposure Prevalence: 100% | OR=1.2 | 1.00 | 0 | 0.0% |
|  | Population-based |  |  | 1.00 | <0.001 | 0.1% |
|  | Pathogen exposed |  | OR=2 | 1.00 | <0.001 | 3.5% |
|  | Population-based |  |  | 1.00 | 0 | 0.0% |

**Table S1**: Heterogeneity estimates across simulated cohorts. Controls indicate whether cases were compared to well-characterized (pathogen exposed) or population-based controls. U1~’Pathogen Exposure’: The scenario-specific parameter for the strength of the relationship between U1 and pathogen exposure. OR: Odds Ratio, reflects the mean OR observed across 1,000 cohorts from each scenario’s parameter-specific simulations. I^2^: Subgroup-specific heterogeneity statistic. Tau(τ)^2^: Subgroup-specific between-study variance component.

**Table S2**: Suggestively associated markers (P<5x10^-5^) from the GWAS comparing HCV clearance cases to ancestry-matched UKB controls.

- [Link to all genome-wide markers](https://my.locuszoom.org/gwas/256899/?token=db1970974d124f2dbc55972db1887a8a), *will subset + upload separate file*

| **Analyzed groups** | **Chromosome 6**  ***HLA-DQB1*, rs9275241** | | | | **Chromosome 19**  ***IFNL3*, rs11881222** | | | |
| --- | --- | --- | --- | --- | --- | --- | --- | --- |
|  | **OR (95% CI)** | **P value** | **Cases (EAF, G)** | **Controls (EAF, G)** | **OR (95% CI)** | **P value** | **Cases (EAF, G)** | **Controls (EAF, G)** |
| Cases vs. well-characterized controls  (HCV Persistence) | 0.61 (0.53-0.71) | 1.90x10^-11^ | 0.39 | 0.51 | 0.42 (0.36-0.50) | 4.06x10^-23^ | 0.20 | 0.36 |
| Cases vs. Ancestry-matched population-based controls (UKB) | 0.69 (0.62-0.77) | 4.35x10^-11^ | 0.39 | 0.52 | 0.61 (0.54-0.69) | 4.04x10^-16^ | 0.20 | 0.28 |
| Cases vs. Ancestry-matched stringent QC population-based controls (UKB) | 0.70 (0.63-0.78) | 2.80x10^-10^ | 0.39 | 0.52 | 0.61 (0.55-0.70) | 5.07x10^-15^ | 0.20 | 0.28 |
| Cases vs. Mahalanobis distance matched UKB controls (1:1) | 0.67 (0.57-0.78) | 8.32x10^-07^ | 0.39 | 050 | 0.50 (0.41-0.6) | 9.58x10^-13^ | 0.20 | 0.32 |
| Cases vs. Mahalanobis distance matched UKB controls (1:10) | 0.68 (0.61-0.77) | 7.16x10^-11^ | 0.39 | 0.49 | 0.59 (0.52-0.68) | 1.27x10^-13^ | 0.20 | 0.29 |
| Cases vs. Propensity score matched UKB controls (1:1) | 0.68 (0.58-0.81) | 4.93x10^-06^ | 0.39 | 0.49 | 0.59 (0.49-0.71) | 2.96x10^-08^ | 0.20 | 0.29 |
| Cases vs. Propensity score matched UKB controls (1:10) | 0.68 (0.61-0.76) | 4.85x10^-11^ | 0.39 | 0.49 | 0.59 (0.51-0.68) | 7.14x10^-14^ | 0.20 | 0.30 |
| Cases vs. Propensity score matched UKB controls (1:10, UKB <2% IBD) | 0.68 (0.61-0.77) | 7.58x10^-11^ | 0.39 | 0.49 | 0.59 (0.51-0.68) | 8.58x10^-14^ | 0.20 | 0.30 |
| Cases vs. Propensity score matched UKB controls  (1:10, No Hemophiliacs) | 0.70 (0.61-0.80) | 1.95x10^-07^ | 0.39 | 0.48 | 0.60 (0.51-0.7) | 5.42x10^-10^ | 0.20 | 0.29 |
| HCV Consortium vs. Ancestry-matched population-based controls (UKB) | 0.92 (0.85-0.99) | 2.54x10^-02^ | 0.47 | 0.52 | 0.96 (0.88-1.04) | 2.98x10^-01^ | 0.30 | 0.28 |

**Table S3**: Variability in effect estimates of known loci across performed GWAS. Marker-specific odds ratios and frequencies of the effect allele are provided for a representative marker selected from each known HCV clearance associated locus. The 95% confidence interval (CI) for the odds ratio and P values observed for each locus-specific marker is provided. IBD: Identity by descent. OR: odds ratio, CI: 95% confidence interval, EAF: effect allele frequency.

| **GWAS** | **Chromosome 2**  ***MIR3681HG*, rs10803744** | | | | **Chromosome 4**  ***STX18*, rs58612183** | | | |
| --- | --- | --- | --- | --- | --- | --- | --- | --- |
|  | **OR (95% CI)** | **P value** | **Cases (EAF, G)** | **Controls (EAF, G)** | **OR (95% CI)** | **P value** | **Cases (EAF, A)** | **Controls (EAF, A)** |
| Cases vs. well-characterized controls  (HCV Persistence) | 1.42 (1.17-1.73) | 4.06x10^-04^ | 0.17 | 0.13 | 1.22 (0.97-1.54) | 8.36x10^-02^ | 0.11 | 0.09 |
| Cases vs. Ancestry-matched population-based controls (UKB) | 1.57 (1.33-1.85) | 7.99x10^-08^ | 0.17 | 0.13 | 1.93 (1.56-2.41) | 3.01x10^-09^ | 0.11 | 0.06 |
| Cases vs. Ancestry-matched stringent QC population-based controls (UKB) | 1.58 (1.34-1.87) | 5.33x10^-08^ | 0.17 | 0.13 | 1.86 (1.5-2.31) | 1.48x10^-08^ | 0.11 | 0.06 |
| Cases vs. Mahalanobis distance matched UKB controls (1:1) | 1.44 (1.14-1.81) | 2.21x10^-03^ | 0.17 | 0.13 | 1.69 (1.27-2.26) | 3.47x10^-04^ | 0.11 | 0.07 |
| Cases vs. Mahalanobis distance matched UKB controls (1:10) | 1.46 (1.26-1.7) | 6.76x10^-07^ | 0.17 | 0.12 | 1.81 (1.5-2.19) | 6.47x10^-10^ | 0.11 | 0.06 |
| Cases vs. Propensity score matched UKB controls (1:1) | 1.76 (1.39-2.24) | 3.85x10^-06^ | 0.17 | 0.11 | 1.69 (1.26-2.26) | 4.08x10^-04^ | 0.11 | 0.07 |
| Cases vs. Propensity score matched UKB controls (1:10) | 1.63 (1.4-1.89) | 2.14x10^-10^ | 0.17 | 0.12 | 1.70 (1.41-2.05) | 2.38x10^-08^ | 0.11 | 0.07 |
| Cases vs. Propensity score matched UKB controls (1:10, UKB <2% IBD) | 1.62 (1.39-1.88) | 3.55x10^-10^ | 0.17 | 0.12 | 1.67 (1.39-2.02) | 5.62x10^-08^ | 0.11 | 0.07 |
| Cases vs. Propensity score matched UKB controls  (1:10, No Hemophiliacs) | 1.52 (1.27-1.82) | 4.09x10^-06^ | 0.18 | 0.12 | 1.47 (1.17-1.84) | 8.55x10^-04^ | 0.10 | 0.07 |
| HCV Consortium vs. Ancestry-matched population-based controls (UKB) | 1.28 (1.14-1.43) | 1.71x10^-05^ | 0.15 | 0.13 | 1.60 (1.38-1.86) | 3.90x10^-10^ | 0.10 | 0.06 |

**Table S4**: Variability in effect estimates of novel loci across performed GWAS. Marker-specific odds ratios and frequency of the effect allele are provided for the top two associated markers from each novel HCV clearance associated locus. The 95% confidence interval for the odds ratio and P values observed for each locus-specific marker is provided. IBD: Identity by descent. OR: odds ratio, CI: 95% confidence interval, EAF: effect allele frequency.

| **Simulation** | **Scenario-specific parameter** | **Fixed Values** | | |  |
| --- | --- | --- | --- | --- | --- |
|  |  | Outcome: Post-exposure | Pathogen Exposure Prevalence | $\beta$:  U1~Pathogen Exposure | Sample size: Cases (N):Controls (N) |
| Scenario 3a: Cases vs. Population-Based Controls | **Outcome: Post-exposure** | **20%,  50%,  80%** | 25% | log(1.2) log(2.0) | 20,000 : 20,000 20,000 : 200,000 |
| Scenario 3b: Cases vs. Well-Characterized Controls |  |  |  |  | 20:000 : 20:000 |

**A**

**B**

|  |  |  |  | 20,000 Cases vs. 20,000 Common Controls | | 20,000 Cases vs. 200,000 Common Controls | |
| --- | --- | --- | --- | --- | --- | --- | --- |
| Scenario | Controls | Parameter Value | U1~Pathogen Exposure | OR (Mean) | Spurious Associations N (%) | OR (Mean) | Spurious Associations  N (%) |
| 3 | Pathogen exposed | Outcome observed: 20% | OR=1.2 | 1 | 0 (0) | 1 | 0 (0) |
|  | Population-based |  |  | 1.04 | 267 (0.05) | 1.04 | 3,720 (0.74) |
|  | Pathogen exposed |  | OR=2 | 1 | 0 (0) | 1 | 0 (0) |
|  | Population-based |  |  | 1.15 | 467,020 (93.4) | 1.15 | 499,984 (>99.99) |
|  | Pathogen exposed | Outcome observed: 50% | OR=1.2 | 1 | 0 (0) | 1 | 0 (0) |
|  | Population-based |  |  | 1.04 | 153 (0.03) | 1.04 | 1,749 (0.35) |
|  | Pathogen exposed |  | OR=2 | 1 | 0 (0) | 1 | 0 (0) |
|  | Population-based |  |  | 1.13 | 407,864 (81.57) | 1.13 | 499,647 (99.93) |
|  | Pathogen exposed | Outcome observed: 80% | OR=1.2 | 1 | 0 (0) | 1 | 0 (0) |
|  | Population-based |  |  | 1.04 | 85 (0.02) | 1.04 | 872 (0.17) |
|  | Pathogen exposed |  | OR=2 | 1 | 0 (0) | 1 | 0 (0) |
|  | Population-based |  |  | 1.12 | 326,038 (65.21) | 1.12 | 496,708 (99.34) |

**Table S5**: Additional simulation scenario (Scenario 3). In Table A, the parameter of interest, the fixed values, and the sample sizes of each simulation is listed. In Table B, the number and percentage of spurious associations and mean odds ratio across the parameter-specific simulations are provided. OR: odds ratio.
