## Supplementary Methods for "Pathogen exposure misclassification can bias association signals in GWAS of infectious diseases when using population-based common controls"

**Simulations to characterize pathogen exposure-associated selection bias**

*Additional simulation scenario (Scenario 3) – Effects of the prevalence of the outcome:* While pathogen prevalence was the main epidemiological parameter of interest, a third simulation scenario was performed to assess whether the proportion of exposed individuals who developed the outcome influenced the observed bias due to differential misclassification of pathogen exposure. We generated 500,000 cohorts of N=1,000,000 individuals, in which 20%, 50%, or 80% of exposed individuals developed the outcome. Simulations were performed assuming a 25% prevalence of pathogen exposure and either a moderate (β=log(1.2), OR=1.2) or a strong association (β=log(2), OR=2) between U1 and pathogen exposure (**Table S5A**).

*Evaluation of heterogeneity of association results.* To determine whether significant heterogeneity across effect estimates were observed within each set of parameter-specific replicates (N=500,000), indicating that we failed to simulate independent but otherwise equivalent cohorts, we performed tests of heterogeneity using 1,000 randomly selected cohorts from each scenario’s parameter-specific simulations using the meta R package.^1^ Based on estimated cohort variance components (τ<0.001) and the proportion of total variation observed across scenario-specific estimates due to heterogeneity (I^2^<10%), there was no significant amount of within-group heterogeneity (**Table S1**).^2–4^

**HCV GWAS using cases and well characterized controls or population-based controls**

*Selection of samples for analysis*

*Stringent QC-passed UKB controls*: To identify the set of UKB controls who passed a more stringent set of sample-level genetic heterozygosity-focused QC metrics than those performed by the UKB consortium, individuals with missingness greater than 3% of all imputed markers from across the genome overlapping with imputed markers from the HCV consortium or had sample-level heterozygosity rates greater than three standard deviations outside the mean were excluded prior to any ancestry-based matching (N=13,986). After ancestry-based matching, 366,354 UKB individuals remained. A second heterozygosity-based filtering step was then performed, which excluded an additional 2,046 UKB controls. A total of 364,308 stringent QC-passed ancestry-matched UKB controls were identified. These sample-level quality control measures were performed using Plink via the plinkQC tool in R.^5^ Marker-level matching with the HCV consortium and QC, as described within the main text, were then performed using this stringent-QC set of UKB controls. A total of 6,009,835 QC-passed imputed markers were shared across the combined HCV Consortium and this set of stringent QC-passed UKB cohort individuals.

To determine whether any other sample-level QC issues could explain the observed novel loci, genotype missingness and allele frequencies for the top associated markers in each of the novel HCV clearance associated were estimated within each of the 106 UKB-defined QC batches limited to the stringent QC-passed UKB control individuals.^6^ The batch-specific allele frequencies for each locus were then compared to the gnomAD-related MAF filter frequency threshold determined for each marker.

*Matched case-control cohorts using different subsets of UKB controls*: Given the extreme case-control imbalance of our HCV clearance individuals compared to the ancestry-matched or stringent QC-passed UKB controls, we were concerned that ancestry-associated PC covariates were driven entirely by UKB controls. We therefore utilized bipartite matching approaches to identify subsets of UKB controls most genetically similar to each HCV clearance individual,^7^ and then performed GWAS with these smaller cohorts of matched controls. For the identification of matched UKB controls at case-control ratios of 1:1 or 1:10, we estimated pairwise distances between each of the 702 individuals with HCV clearance and all stringent QC-passed UKB individuals using either a Mahalanobis or propensity score discrepancy-based distance. Mahalanobis distances were derived using the top ten PCs while propensity score-based distances were derived using the top ten PCs and sex. Distance estimates and subsequent bipartite matching at the above ratios of cases to controls were performed using each distance metric with the optmatch package in R.^7^ The PCs utilized for distance estimation were derived from the combined HCV consortium and stringent QC-passed UKB controls. For the 1:1 matching, an average of one control per case was successfully obtained and 702 cases were compared to 702 UKB controls for each distance-based matching scenario. As 5% (N=36) of the HCV clearance individuals had <5 potential UKB matches, the obtained matched case-control cohorts had ratios slightly below the desired 1:10 ratios. For the Mahalanobis distance-based matching approach, a total of 6,633 UKB controls were identified in the 1:10 case-control ratio scheme. For the propensity score-based matching approach, a total of 6,666 UKB controls were identified in the 1:10 case-control ratio scheme.

*Variability of estimated effects across alternative matched case-control cohorts:*

To explore the extent to which observed effect estimates vary across alternative yet plausible matched case-control cohorts, we performed 1,000 additional 1:10 case-control matches using each distance metric. This involved 1,000 random selections of ten UKB controls for each of the 702 HCV cases, where possible. This resulted in 2,000 alternative matched case-control cohorts. The same covariates utilized in each GWAS (sex and the top 20 PCs) were included in the genetic associations performed when using the 2,000 alternative sets of matched case/population-control individuals. PCs used as covariates were estimated from the combined HCV Consortium and stringent QC-passed UKB controls.

*Exploratory GWAS – excluding individuals with hemophilia*: Hemophilia-based exclusion was accomplished by excluding cases from any HCV consortium sub-study which used hemophilia as an inclusion criterion and their propensity score matched controls (1:10). It is possible that some remaining cases included in this comparison also had hemophilia, but we were not able to exclude them since it was not phenotypically characterized and/or annotated.

*Exploratory GWAS – minimal IBD sharing UKB controls*: For the identification of pairs of UKB individuals who share identity-by-descent (IBD) proportions under 2%, we used the KING toolset.^8^ Relatedness and shared IBD proportions were estimated for the 6,666 propensity score-based matched UKB controls (1:10 ratio), identifying 217 pairs of UKB individuals who shared IBD proportions >2%. After excluding 154 of these UKB controls, a total of 6,512 matched UKB controls were included in the IBD-filtered GWAS, in which no pair of UKB controls had >2% shared IBD.

***Statistical analysis***

**HCV GWAS using cases and well characterized controls or population-based controls**

*Exploratory GWAS – Genetic Associations:*

In addition to the main GWAS performed comparing 702 HCV clearance cases to 370,702 ancestry-matched UKB controls, REGENIE was used for the GWAS comparing HCV clearance cases to the stringent QC-passed UKB controls. The whole genome penalized regression step of REGENIE was fit using pruned markers for each GWAS (N~90,000 markers), while all QC-passed imputed markers were included in logistic regression score-based association testing. As extreme case-control imbalances were not an issue for the matched case-control exploratory GWAS, these GWAS were performed via logistic regression in Plink.^9,10^ For all GWAS, sex and the top 20 PCs were included as covariates.

*Exploratory GWAS – Heterogeneity of effect estimates:*

To assess whether the observed effect estimates differed between the cases/population-based controls and cases/stringent QC passed population-based controls we performed Cochran’s *Q* test of heterogeneity using METAL.

*Variability of estimated effects across* *alternative matched case-control cohorts:*

We performed multiple logistic regressions with the speedglm package in R using the genotypes of the top associated markers across four loci of interest across the 2,000 alternatively matched case-control cohorts. In addition to visual comparisons of the resulting effect estimate distribution relative to estimates obtained from our main GWAS, we compared the variability in effect estimates across each locus by estimating the coefficient of variation (CoV) which partially accounts for the difference in MAFs, thereby allowing the comparison of variability estimates between loci of different MAFs. We also performed two-way analysis of variance tests to determine whether matching ratio or distance metric was significantly associated with the variation in marker-specific effect estimates and Anderson-Darling k-Sample tests to determine whether the use of different matching approaches or ratios resulted in significantly different distributions of marker-specific effect estimates.^11^

***Supplementary Results:***

**Simulations to characterize pathogen exposure-associated selection bias**

*Additional simulation scenario (Scenario 3) – Effects of the prevalence of the outcome:* In this scenario, inflated ‘outcome of interest’~SNP effect estimates were consistently observed, regardless of the number of population-based controls used and the prevalence of the outcome among controls (**Table S5 B**). This bias was negatively associated with the outcome-specific parameters (P=0.03), indicating that the higher the prevalence of the outcome among population-based common controls, the lower the proportion of spurious associations **(Figures S15-S16)**.

**HCV GWAS using cases and well characterized controls or population-based controls**

*Exploratory GWAS – Heterogeneity of effect estimates:*

No significantly or suggestively associated marker shared across these cohorts was observed to have a significantly different effect size between the stringent QC-passed UKB controls GWAS compared to the ancestry-matched population-based UKB controls GWAS.

*Variability of estimated effects across alternative matched case-control cohorts:* We observed significant variability in the effect estimates of both known and novel loci in the 2,000 alternative matched case-control cohorts **(Figure S17)**. The distribution of observed effect sizes for the top two markers from each novel loci were more variable (CoV*_MIR3681HG_*=6.96, CoV*_STX18_*=6.64) compared to a representative single marker from each of the known loci (CoV*_HLA-DQB1_*=3.89, CoV*_IFNL3_*=5.05). For all loci, analysis of variance tests indicated that the choice of distance metric (e.g., Mahalanobis distance vs. propensity score-based distance) was significantly associated with the variability in effect estimates (P<0.0001). For each locus, multiple distinct distributions (>1) were observed in the Anderson-Darling k-Sample tests when the matching scenarios were compared simultaneously (P<0.0001). These results indicate the method by which exact case-control matching is accomplished in a GWAS can have significant ramifications on the resulting effect estimates. For example, several of our regression-based matching scenarios resulted in the *IFNL3* locus being only suggestively associated with HCV clearance **(Figure S17)**. We should note that the extreme deflation in the effect estimates for *IFNL3* observed in the main GWAS was not present when ancestry-associated PCs were re-estimated in each matched case-control scenario. Indeed, for each matched case-control GWAS in which the top twenty PCs were re-estimated and included as covariates, the observed effect size estimates for the known loci were similar to those observed in the HCV clearance vs. ancestry-matched UKB controls GWAS results (**Table S3**).

In addition to findings related to the appropriateness of shared controls for GWAS of infectious diseases related outcomes, the highly variable GWAS results observed across our alternative matched case-control cohorts highlights the importance specific bipartite matching approaches have upon any eventual GWAS findings. These results also demonstrate the importance of re-estimating PCs for each GWAS, as genetic associations performed using globally estimated PCs were highly biased compared to the results obtained when using alternative matched case-controls cohort derived PCs.

*Exploratory analysis – Novel HCV Clearance-associated loci:*

We observed significant variability in the allele frequency for the top two HCV clearance associated *STX18* markers across the 106 UKB-defined QC batches (rs3935096: 5.58%-7.19%, rs58612183: 5.5%-7.16%),^6^ with 6.6% of batches failing our gnomAD allele frequency-based QC threshold. As the average MAF across all UKB batches did not fall outside the gnomAD-derived MAF threshold for either of the top two HCV clearance associated markers within *STX18*, this locus was retained.

We found that association analysis of certain alternative matched case/control cohorts obtained with the bipartite matching approaches resulted in significantly less extreme effect sizes for the novel loci. We also observed that the matching method was another source of variability; for example, the 1:1 Mahalanobis distance-based matched GWAS resulted in the deflation of the *MIR3681HG* signal to values approximating the HCV clearance vs. HCV persistence GWAS estimates (rs10803744 with OR=1.44) and the 1:1 propensity score-based matched GWAS resulted in inflated effect estimates for the *MIR3681HG* locus which were larger than the GWAS involving all ancestry-matched UKB controls (rs10803744 with OR=1.76) (**Table S4**). Future work should determine whether certain distance-based matching schemes are more appropriate for subsampling-based efforts to identify matched common controls from biobank-scale cohorts.

*Previous associations of the HCV novel loci described in former GWAS using population-based controls:* Most previously described associations within *STX18* have involved GWAS performed using population-based controls from different sources/biobanks. These associations have included: congenital heart disease (UKB and Wellcome Trust Case Control Consortium/TwinsUK controls),^12^ acute myeloid leukemia (HapMap controls),^13^ and multiple allergic diseases (>50% of controls from UKB)^14^ as well as suggestive associations with COVID-19 susceptibility (UKB controls) and COVID-19-associated pneumonia (23andMe controls).^15,16^ Similarly, UKB-specific signals for *MIR3681HG* include associations with total/bioavailable testosterone levels in UKB women of European ancestry,^17^ autoimmune thyroid disease when using European UKB controls,^18^ medication use among European UKB participants,^19^ body mass index (BMI) among European UKB participants,^20^ height of all UKB participants,^21^ and educational attainment in a meta-analysis involving UKB controls.^22^ Similar to the *MIR3681HG* COVID-19 association, another locus with a similar significance pattern was observed in *LINGO2* which was genome-wide significant only in the analysis of a single COVID-19 UKB data freeze and not present in further analyses despite increasing case numbers in later freezes. This locus was suggestively associated with HCV clearance when using ancestry matched UKB controls (top marker within *LINGO2* was rs10123736, OR=1.34, P=**7.62x10^-6^) but not in the HCV clearance vs. persistence GWAS (OR=1.36, P=1.29x10^-4^).**

***References***

1. Balduzzi, S., Rücker, G., and Schwarzer, G. (2019). How to perform a meta-analysis with R: a practical tutorial. Evid. Based. Ment. Health *22*, 153–160.

2. Higgins, J.P.T., and Thompson, S.G. (2002). Quantifying heterogeneity in a meta-analysis. Stat. Med. *21*, 1539–1558.

3. Viechtbauer, W. (2005). Bias and Efficiency of Meta-Analytic Variance Estimators in the Random-Effects Model. J. Educ. Behav. Stat. *30*, 261–293.

4. Schwarzer, G., Carpenter, J.R., and Rücker, G. (2015). Meta-Analysis with R (Cham: Springer International Publishing).

5. Meyer, H. V (2021). plinkQC: Genotype quality control in genetic association studies.

6. Bycroft, C., Freeman, C., Petkova, D., Band, G., Elliott, L.T., Sharp, K., Motyer, A., Vukcevic, D., Delaneau, O., O’Connell, J., et al. (2018). The UK Biobank resource with deep phenotyping and genomic data. Nature *562*, 203–209.

7. Hansen, B.B., and Klopfer, S.O. (2006). Optimal Full Matching and Related Designs via Network Flows. J. Comput. Graph. Stat. *15*, 609–627.

8. Manichaikul, A., Mychaleckyj, J.C., Rich, S.S., Daly, K., Sale, M., and Chen, W.-M. (2010). Robust relationship inference in genome-wide association studies. Bioinformatics *26*, 2867–2873.

9. Chang, C.C., Chow, C.C., Tellier, L.C., Vattikuti, S., Purcell, S.M., and Lee, J.J. (2015). Second-generation PLINK: rising to the challenge of larger and richer datasets. Gigascience *4*, 7.

10. Purcell, S., Neale, B., Todd-Brown, K., Thomas, L., Ferreira, M.A.R., Bender, D., Maller, J., Sklar, P., De Bakker, P.I.W., Daly, M.J., et al. (2007). PLINK: A tool set for whole-genome association and population-based linkage analyses. Am. J. Hum. Genet. *81*, 559–575.

11. Baumgartner, W., WeiB, P., and Schindler, H. (1998). A Nonparametric Test for the General Two-Sample Problem. Biometrics *54*, 1129.

12. Cordell, H.J., Bentham, J., Topf, A., Zelenika, D., Heath, S., Mamasoula, C., Cosgrove, C., Blue, G., Granados-Riveron, J., Setchfield, K., et al. (2013). Genome-wide association study of multiple congenital heart disease phenotypes identifies a susceptibility locus for atrial septal defect at chromosome 4p16. Nat. Genet. *45*, 822–824.

13. Lv, H., Zhang, M., Shang, Z., Li, J., Zhang, S., Lian, D., and Zhang, R. (2017). Genome-wide haplotype association study identify the FGFR2 gene as a risk gene for Acute Myeloid Leukemia. Oncotarget *8*, 7891–7899.

14. Ferreira, M.A., Vonk, J.M., Baurecht, H., Marenholz, I., Tian, C., Hoffman, J.D., Helmer, Q., Tillander, A., Ullemar, V., van Dongen, J., et al. (2017). Shared genetic origin of asthma, hay fever and eczema elucidates allergic disease biology. Nat. Genet. *49*, 1752–1757.

15. Shelton, J.F., Shastri, A.J., Ye, C., Weldon, C.H., Filshtein-Sonmez, T., Coker, D., Symons, A., Esparza-Gordillo, J., Chubb, A., Fitch, A., et al. (2021). Trans-ancestry analysis reveals genetic and nongenetic associations with COVID-19 susceptibility and severity. Nat. Genet. *53*, 801–808.

16. Florian Thibord, Melissa V. Chan, Ming-Huei Chen, A.D.J. (2021). A year of Covid-19 GWAS results from the GRASP portal reveals potential SARS-CoV-2 modifiers v2. MedRxiv.

17. Ruth, K.S., Day, F.R., Tyrrell, J., Thompson, D.J., Wood, A.R., Mahajan, A., Beaumont, R.N., Wittemans, L., Martin, S., Busch, A.S., et al. (2020). Using human genetics to understand the disease impacts of testosterone in men and women. Nat. Med. *26*, 252–258.

18. Saevarsdottir, S., Olafsdottir, T.A., Ivarsdottir, E. V., Halldorsson, G.H., Gunnarsdottir, K., Sigurdsson, A., Johannesson, A., Sigurdsson, J.K., Juliusdottir, T., Lund, S.H., et al. (2020). FLT3 stop mutation increases FLT3 ligand level and risk of autoimmune thyroid disease. Nature *584*, 619–623.

19. Wu, Y., Byrne, E.M., Zheng, Z., Kemper, K.E., Yengo, L., Mallett, A.J., Yang, J., Visscher, P.M., and Wray, N.R. (2019). Genome-wide association study of medication-use and associated disease in the UK Biobank. Nat. Commun. *10*, 1891.

20. Zhu, Z., Guo, Y., Shi, H., Liu, C.-L., Panganiban, R.A., Chung, W., O’Connor, L.J., Himes, B.E., Gazal, S., Hasegawa, K., et al. (2020). Shared genetic and experimental links between obesity-related traits and asthma subtypes in UK Biobank. J. Allergy Clin. Immunol. *145*, 537–549.

21. Kichaev, G., Bhatia, G., Loh, P.-R., Gazal, S., Burch, K., Freund, M.K., Schoech, A., Pasaniuc, B., and Price, A.L. (2019). Leveraging Polygenic Functional Enrichment to Improve GWAS Power. Am. J. Hum. Genet. *104*, 65–75.

22. Lee, J.J., Wedow, R., Okbay, A., Kong, E., Maghzian, O., Zacher, M., Nguyen-Viet, T.A., Bowers, P., Sidorenko, J., Karlsson Linnér, R., et al. (2018). Gene discovery and polygenic prediction from a genome-wide association study of educational attainment in 1.1 million individuals. Nat. Genet. *50*, 1112–1121.
